# Genome-wide association study meta-analysis brings monogenic hearing loss genes into the polygenic realm

**DOI:** 10.1101/2025.02.28.25323059

**Authors:** Royce Clifford, Jacquelyn Johnson, Caroline E Mackey, Elizabeth A Mikita, Allen F Ryan, Million Veteran Program, Adam X Maihofer, Caroline M Nievergelt

## Abstract

**Background:** Disabling age-related and noise-induced sensorineural hearing loss (SNHL) affects 5% of the global population and is associated with isolation, depression, cognitive decline, and dementia. SNHL is strongly heritable with a polygenic signature of common genetic variants conferring small risks individually, thus requiring very large studies to achieve statistical power in genome-wide association studies (GWAS).

**Methods:** We present the first report of a clinical SNHL GWAS in the Million Veteran Program (MVP) cohort. GWAS findings are examined separately within MVP (210,240 cases and 265,275 controls), including multi-ancestry analysis, then combined and contrasted with the United Kingdom Biobank (UKB) self-reported hearing loss study (87,056 cases and 163,333 controls). We perform functional mapping and annotation, gene prioritization, gene-based and gene-set analysis, and cochlear cell type enrichment, including human single-cell results, then compare our results to known hereditary hearing loss genes. Summary results are leveraged to characterize the genetic architecture of SNHL.

**Results:** Substantial genetic overlap is seen between MVP and UKB despite differences in phenotypes, demographics, and environmental exposures. Individual GWAS and meta-analyses identify 108 loci, including 54 loci containing novel prioritized genes and/or protein-coding genes. Significant gene ontology pathways include “sensory perception of mechanical stimulus,” “ear development,” “actin-binding,” and “cytoskeletal protein binding,” indicative of mechanosensory structure and function in the cochlea. GWAS and gene-based analyses implicate 17 known familial hearing loss genes, expressing proteins in regions of hair cells, Type 1 neurons, stria vascularis, and subcellular regions of stereociliae, rootlets, and notably, mechanoelectrical transduction tip-links. Although risk for polygenic SNHL is predominantly captured by SNPs outside of congenital HHL genes (HHL), SNP-based partitioned heritability estimates show a 3.26-fold enrichment of HHL relative to other genes.

**Conclusion:** In the largest GWAS to date, combining clinical diagnosis of hearing loss and self-report data from two cohorts, we identify 108 loci with 54 novel genes. Despite the increased enrichment of HHL genes, 97% of the risk for adult SNHL is captured by SNPs outside of HHL genes. Although SNHL in the UKB and MVP were assessed using different phenotypes, genetic signals between the two cohorts are predominantly shared, and locus discovery is boosted through increased sample size in meta-analysis.

## Introduction

The effects of permanent sensorineural hearing loss (SNHL) range from annoyance at a partner’s “mumbling” to a disabling disorder that currently affects 1.6 billion people globally, a number that is predicted to increase by greater than 50% over the next three decades^1^. On a personal level, SNHL is associated with isolation, depression, cognitive decline, frailty and dementia^2–4^.

The most common causes of SNHL are aging and noise exposure^5^. Twin-studies have calculated a heritability for SNHL of 0.35-0.65^6,7^, indicating that a large portion is accounted for by genetic factors. Although early-onset hearing impairment has been linked to roughly 200 human hereditary hearing loss (HHL) genes,^8^ in contrast, findings from genome-wide association studies (GWAS) on adult onset SNHL demonstrate a highly polygenic genetic architecture^9,10^, characterized by common genetic variants individually conferring small risks.

Whereas the technology to cure hearing impairment caused by monogenic disorders utilizing gene therapy is rapidly advancing with some syndromes currently undergoing clinical trials^11^, ultimate treatment of the more common polygenic forms of SNHL will require identification and characterization of multiple genes, variants, and pathways using technology yet to be determined. Given that statistical power to identify risk loci for polygenic disorders mandates large genetic studies, much of the otologic research has focused on the use of one or two self-reported items for disease identification^10,12^. While the audiogram remains the gold standard for an objective measure of hearing loss, self-reported hearing loss questions have provided a satisfactory measure to elicit relevant loci from large genome-wide association studies (GWAS)^13–15^. The largest self-reported SNHL GWAS meta-analysis to date, including the UK Biobank (UKB) and 8 smaller cohorts of clinically diagnosed and self-reported hearing impairment on 723,266 individuals from the EARGEN consortium, identified 48 loci^16^.

Here we report on a phenotype based on ICDs (international classification of disease) that include the most common forms of SNHL, i.e., age-related and noise-induced, typically ascertained by audiograms, from the Million Veteran Program (MVP), comparing the limitations of measuring a complex and clinically heterogeneous trait based on self-report to a clinical diagnosis of SNHL derived from more precise auditory measurements.

MVP consists of an aging population currently with over one million participants, who, during their military career, have been exposed to multiple sources of loud noise^17^. To address the question of ’minimal phenotyping’, we contrast the GWAS based on clinical diagnoses from MVP with a GWAS on self-reported hearing loss in the United Kingdom Biobank (UKB), which has been included in most of the recent GWAS meta-analyses^10,12,16,18–22^. Our findings indicate that despite the differences of phenotype, demographics, and noise exposure of the two cohorts, they share a large proportion of their genetic architecture. Combination in meta-analysis elicits a total of 108 loci in individual ancestry GWAS and meta-analysis with 54 novel genes from prioritized analysis and 100 significant novel genes from gene-based examination. These loci contain inferences of 17 congenital hearing loss genes as well as multiple other ear-related genes, and we present a plausible explanation for a mechanism of injury.

## Methods

### SNHL Phenotype in MVP

MVP provided data from 963,753 participants (658,038 with genotype data) in version 23_1 (released Apr 29, 2024), recruited 2011 through Sep 30, 2023, consisting of ICD records from the US Veterans Administration Corporate Data Warehouse^23^. Information was linked to individual, de-identified electronic health records (EHR).

Hearing loss was assessed using ICD9/ICD10 codes; participants with the following codes were excluded from analysis: otosclerosis (H80.0, H80.1, H80.2, H80.8, H80.9, 387.0, 398.1, 387.2, 387.8, and 387.9), Meniere’s disease (H81.0 and 386.0), pulsatile and objective tinnitus (H93.A, 388.32), conductive hearing loss (H90.0, H90.1, H90.2, H90.A1, 389.0), mixed hearing loss (H90.6, H90.7, H90.8, H90.A, H90.A3, 389.2), unilateral sensorineural hearing loss (H90.4, H90.A2, 389.13, 389.15, 389.17), ototoxicity (H91.0), sudden hearing loss (H91.2, 388.2) congenital malformations and anomalies (Q16, Q17.2, Q17.4, Q17.8, Q17.9, 744.0, 733.23, 744.29), intraoperative and postprocedural complications and disorders of ear and mastoid process, including mastoiditis and related conditions (H95, 383.3, 383.8, 383.89), certain degenerative and vascular disorders of the ear (H93.0, 388.0, 388.00, 388.02), disorders of the acoustic and cranial nerves (D33.3, H93.3, 225.1, 388.5), acoustic neuritis in infectious and parasitic diseases (H94.0), in addition to Deaf non-speaking and other specified forms of hearing loss (H91.3, H91.8, H94.8, 389.7, 389.8).

From the remaining participants, SNHL cases (N = 236,597) had to have a diagnosis of sensorineural hearing loss (H90.3, H90.5, H91.1, H91.10, H91.11, H91.12, H91.13, 388.01, 389.1, 389.10, 389.11, 389.12, 389.14, 389.16, and 389.18), while controls (N = 296,284) consisted of those who had no evidence of the above ICDs nor ICDs for unspecified hearing loss (H91.9, H91.90. H91.91, H91.92, H91.93), or for the fitting, adjustment, presence, or management of a hearing aid or device (Z45.3, Z46.1, Z96.2, Z97.4, 389.9, and V53.2). All participants provided written informed consent, and the study was approved by the University of California San Diego and VA Central Institutional Review Boards.

### Genotyping, quality control, and imputation

Analyses were based on release 4 of the MVP imputed genotype data containing 658,038 participants. Details of the genotyping, quality control, and imputation procedures used have been reported in detail^23^. In brief, MVP samples were genotyped on a customized version of the Affymetrix Axiom biobank array and standard genotype quality control procedures were followed. Genotype data was phased using Eagle version 2.4^24^ and imputed using minimac version 4^25^ with the Haplotype Reference Consortium^26^ reference panel.

### Assessment of ancestry

Ancestry was determined using SNPweights^27^ with a global reference panel^28^, including 93,172 SNPs that overlap between the reference data and MVP genotype data. Reference SNPs were selected to have MAF > 1% in reference populations and are in low LD (based on independent pairwise LD pruning, using a 1000 kb window, 50 SNP step size, and r-squared of 0.2, as calculated in PLINK^29^. Participants were placed into the three following large groupings: 1. European ancestry (EUA; individuals with ≥ 90% European ancestry); 2. African ancestry (AFA; individuals with ≥ 5% African ancestry, < 90% European ancestry, < 5% East Asian, Native American, Oceanian and Central-South Asian ancestry, or individuals with ≥ 50% African ancestry, < 5% Native American, Oceanian and < 1% Asian ancestry); and 3. Indigenous American Ancestry (IAA; individuals with ≥ 5% Native American ancestry, < 90% European, and < 5% African, East Asian, Oceanian and Central-South Asian ancestry).

### Calculation of relatedness and principal components (PCs)

Relatedness within samples was estimated using KING^30^. For each pair of related participants (kinship coefficient > 0.0884), one was removed, giving preference to retain cases. If case status was identical between the pair, one of the pair was removed at random (N = 21,923 excluded). PCs were calculated within unrelated subjects of the same ancestry using FlashPCA2^31^. SNPs were excluded for MAF < 1%. Remaining SNPs were pruned for LD over 1 MB windows stepped over 50 variants at a time with an r^2^ threshold of 0.05.

### GWAS in MVP based on EHR data

GWAS for SNHL was performed for each of the 3 ancestry groups in MVP separately with PLINK 2^29^, using logistic regression including sex, age, age^2^, and 10 PCs as covariates. Only SNPs with MAF > 1% and imputation info score > 0.6 (calculated within analytic sample) were analyzed. Genome-wide significance (GWS) was assessed at a *p* value < 5×10^-8^.

### GWAS in UK Biobank (UKB) based on self-reported hearing difficulty

GWAS summary results from Wells et al.^12^ for self-reported hearing difficulty in the UKB were downloaded from the UKB Returned Datasets Catalog. The cohort consisted of white British ancestry (EUA only) recruited from 2007 - 2013 who volunteered to participate and completed a questionnaire including field code 2247 and 2257 regarding hearing difficulties and difficulties hearing in ambient noise. GWAS cases were those who answered “yes” to both questions ("Yes, diagnosed by doctor or health professional’’ or ‘‘Yes, not diagnosed by health professional’’; N = 87,056 cases), controls were those who answered “no” to both (N = 163,333 controls). Association analysis was performed using a linear mixed-effects model implemented in BOLT-LMM v.228 including age, sex, genotyping platform, and 10 PCs (see ^12^ for additional details).

### Meta-analysis

For compatibility with MVP, UKB GWAS summary statistics were converted to the log odds ratio scale^32^ and inverse variance weighted fixed effects meta-analysis was performed in METAL^33^. Only SNPs present in two or more input datasets were included in meta-analyses. Three meta-analyses were performed in total: 1. MVP multi-ancestry (EUA, AFA, IAA) meta-analysis; 2. European ancestry meta-analysis in MVP and UKB; 3. meta-analysis of the full sample, including MVP multi-ancestry + UKB EUA GWAS.

### Functional mapping and annotation

Functional annotation of the GWAS results were performed with FUMA v1.6.1^34^ using default settings and the human genome assembly GRCh37 (hg19). The SNP2Gene module was used to define independent genomic risk loci and variants in LD with lead SNPs (r^2^ > 0.6, calculated using ancestry appropriate 1KGPp3 reference genotypes (EUR, AMR, AFR). Genomic risk loci were combined to have non-overlapping base pair positions and SNPs were annotated to the nearest gene. Functional consequences of SNPs were obtained by mapping SNPs on their chromosomal position and reference alleles to databases containing known functional annotations, including ANNOVAR^35^, Combined Annotation Dependent Depletion^36^ and RegulomeDB^37^.

### Gene prioritization

Gene prioritization was performed using FLAMES version 1.1.2^38^. To generate credible sets, Bayesian fine-mapping of risk loci was performed using the coloc R package^39^. Convergence-based gene prioritization scores were calculated using Polygenic Priority Scores v0.2^40^, using as inputs MAGMA gene-level z-scores and pathway naïve scores. FLAMES annotations were generated for each credible set, using as inputs the functional annotation data, Polygenic Priority Score output data, and FUMA generated MAGMA gene and gene-tissue analysis. Note, while FUMA positionally maps SNPs to genes based on their proximity to gene boundaries (within 10kb), FLAMES annotates risk loci to a potentially larger set of genes because, in addition to positional mapping, it incorporates several other SNP-to-gene annotation databases. FLAMES scoring was performed using the annotation data. Only genes above the previously calibrated cumulative 75% precision threshold are reported.

### Hearing genes from HereditaryHearingLoss.org

We extracted 196 protein-coding genes associated with non-syndromic and syndromic hearing loss from the Hereditary Hearing Loss database (https://hereditaryhearingloss.org, accessed 8/9/2024)^8^. This excluded Mitochondrial Non-syndromic Hearing Loss genes and *MIR96,* which encodes microRNA. Protein-coding genes were aligned with genes from GWS genomic risk loci and gene-based analysis (MAGMA). Genes with multiple symbol aliases were consolidated using the NIH gene database (https://www.ncbi.nlm.nih.gov/gene/ ; accessed 8/28/2024).

### Identification of novel genes

In order to identify novel, previously not reported genes, the following criteria were used. For GWS loci from the GWAS: 1. The locus did not have any genes that were present in the HereditaryHearingLoss.org database (accessed 8/9/2024), 2. the locus did not have any genes that were present in the NHGRI-EBI GWAS Catalog database (https://www.ebi.ac.uk/gwas/home, accessed 7/2/2025; selected traits: sensory perception of sound, hearing loss, sensorineural hearing loss, hearing threshold trait, sensorineural hearing impairment, hearing physiology trait, noise-induced hearing loss, age-related hearing impairment, hearing process quality, able to hear with hearing aids, deafness, but excluding chemoradiation-induced hearing loss in nasopharyngeal carcinoma and speech-in-noise perception traits, as well as SNP-by-SNP and SNP-by-environment interaction studies and intergenic loci). 3. the locus does not overlap with a GWS locus from Wells 2019^12^ hearing difficulty (since this GWAS was used for the meta-analysis), 4. the locus did not overlap with something that was disqualified due to #1, 2, or 3, and 5. the locus had at least one protein-coding gene in the locus and/or a FLAMES prioritized gene.

For genes from the gene-based analysis: points 1-3 were applied.

### Regional association plots

Regional visualizations of GWS loci were produced using LocusZoom 1.4^41^. Linkage Disequilibrium (LD) was calculated using 1KGPp3 data, using ancestry-specific reference data (EUR, AMR, AFR).

### Gene-based and gene set analyses with MAGMA

The Multi-marker Analysis of GenoMic Annotation (MAGMA) v1.08^42^ tool implemented in FUMA was used to perform gene-based, gene-pathway, and human-based tissue enrichment analyses. For gene-based analysis, SNPs were mapped to 18,809 protein coding genes. For each gene, its association with SNHL was determined as the weighted mean squared test statistic of SNPs mapped to the gene, where LD patterns were calculated using ancestry appropriate 1KGPp3 reference genotypes. The significance of genes was set at a Bonferroni-corrected threshold of p < 2.658e-06 (0.05/18,809). To identify specific biological pathways implicated, gene-based test statistics were used to perform a competitive set-based analysis of 17,012 pre-defined curated gene sets and GO terms obtained from MsigDB^43^. The significance of pathways was set at a Bonferroni-corrected threshold of p < 2.94e-06 (0.05/17,012).

### Cochlear cell type enrichment analyses in human

To test if tissue-specific gene expression was associated with SNHL, enrichment analyses were performed via MAGMA gene-tissue analysis of the EUA gene-based GWAS results. Since human inner ear cell type data has not yet been integrated in the FUMA tissue collection, we paired the collection with human adult inner ear tissue ^44^. Data was converted from h5ad to h5seurat using SeuratDisk (0.0.0.9021). The R package Seurat v5.3.0 function AggregateExpression() was used to log normalize and return pseudobulk gene expression across each cell type. MAGMA v1.10 was used to annotate the SNPs based on GRCh37 and perform gene-set analysis with GTEx v8 specific and general gene expression datasets downloaded from FUMA.^34^ Gene-set analyses were conditioned on the mean of the normalized gene expressions (reads per kilo base per million, RPKM) of the GTEx v8 tissues.

### Cochlear cell type enrichment analyses in mice

Cochlear cell type enrichment analyses were performed via MAGMA gene-tissue analysis of the EUA gene-based GWAS results paired with mice expression data. Mouse gene expression data was derived from cochlear cells and nuclei from Jean *et al.* (20-day old mice only)^45^ and Hoa et al^46^. Additional pre-processing steps were applied to data from Jean *et al.*; cells were retained if the RNA count was between 1,000 and 30,000, gene/non-coding RNA between 500 and 5,000, and mitochondrial genes less than 2% using the R package Seurat v4.9.9.9058. Pseudobulk gene expression was calculated by log2 normalizing the mean raw expression across each cell type. Data from Hoa et al. for the 5 tissues from the organ of Corti had been pre-processed into reads per kilobase million by study authors. Mouse gene Ensembl IDs were converted to human Entrez IDs using a homology map from the Mouse Genome Database^47^. Only genes with one-to-one mappings were kept. MAGMA v1.10 was used to annotate the SNPs based on GRCh37 and perform gene-set analysis. Gene-set analyses were conditioned on the log2 mean expression across the cochlear cell types for each gene.

### SNP-based heritability and genetic correlation

SNP-based heritability (h^2^_SNP_) was evaluated in EUA data using LD score regression (LDSC)^48^. Input LD scores were computed from 1KGPp3 EUA samples. The LDSC intercept was used to test for artifactual inflation of test statistics, and the attenuation factor was used to estimate the proportion of inflation coming from polygenic signal. The h^2^_SNP_ was converted to the liability scale^49^. Cross-trait LDSC^48^ was used to estimate the genetic correlation (r_g_) between datasets.

### Univariate and bi-variate Gaussian mixer model (MiXeR) analysis

We used univariate MiXeR v1.3^50^ to estimate the genetic architecture of MVP EUA and UKB SNHL phenotypes. MiXeR estimates SNP-based heritability and two subcomponents whose product is proportional to heritability: the proportion of non-null SNPs (polygenicity) and variance of effect sizes of non-null SNPs (discoverability). MiXeR was applied to GWAS summary statistics under the default settings with the supplied EUA LD reference panel. The results for the number of influential variants reflect the number of SNPs necessary to explain 90% of SNP based heritability. Bivariate MiXeR^51^ was used to estimate phenotype specific polygenicity and the shared polygenicity between phenotypes. Goodness-of-fit of the MiXeR model relative to simpler models of polygenic overlap was assessed using AIC values. Heritability, polygenicity and discoverability estimates were contrasted between datasets using the *z*-test.

## Results

### Ancestry-specific GWAS and multi-ancestry meta-analysis for SNHL in MVP

GWAS in the MVP was performed based on ICD diagnosis of SNHL from the electronic health record. After excluding participants for mixed, conductive, and uncommon forms of SNHL (e.g. Meniere’s disease and ototoxicity), 210,240 cases and 265,275 controls were available for GWAS. The MVP cohort is predominantly (91%) male with an average age of 62.2 (13.9 SD) years at enrollment (Supplementary Table 1). GWAS were stratified into three ancestry groups. GWAS of European ancestry participants (MVP EUA; 176,393 cases and 172,117 controls) identified 43 genome-wide significant (GWS) loci (Supplementary Table 2, Fig. 1a). The inflation of test statistics (Genomic control λ = 1.33) was almost entirely accounted for by polygenic effects (LDSC attenuation ratio: 14.7%; LDSC intercept = 1.06; SE = 0.009; Supplementary Table 7). Protein-coding genes within GWS loci were identified using functional annotation with FUMA and further prioritized using FLAMES, identifying 29 genes. GWAS of African ancestry participants (MVP AFA; 22,757 cases and 74,842 controls) identified one GWS locus (rs59174224, p = 4.12E-08) within the gene *TUB* (Fig. S1b. *Tub* is a known murine gene leading to profound hearing loss. A modifier *Moth1* protects hearing, and *Mtap1* then mediates *Moth1* at the synaptic terminal^52^. GWAS of MVP indigenous American ancestry participants (MVP IAA; 11,090 cases, 18,316 controls) did not identify any GWS loci (Fig. S1c). Meta-analysis of the EUA, AFA, and IAA GWAS (MVP meta-analysis; N = 475,515 subjects) identified 52 loci; 14 of which were not significant in individual ancestry analyses (Supplementary Table 2, Fig. S1d). Gene prioritization using FLAMES identified 36 genes in the 52 loci. Annotations for SNPs in LD with significant SNPs in the MVP multi-ancestry meta-analysis are summarized in Supplementary Table 3. Of the 52 loci, 12 contained a GWS SNP with a CADD score predicted to be deleterious to function (>12.37) that was within the exon region of a gene. A total of 57 independent GWS loci were identified across the four MVP GWAS performed. Regional association plots comparing ancestry-specific analyses for these loci are shown in Supplementary Data 1.

**Figure 1.**
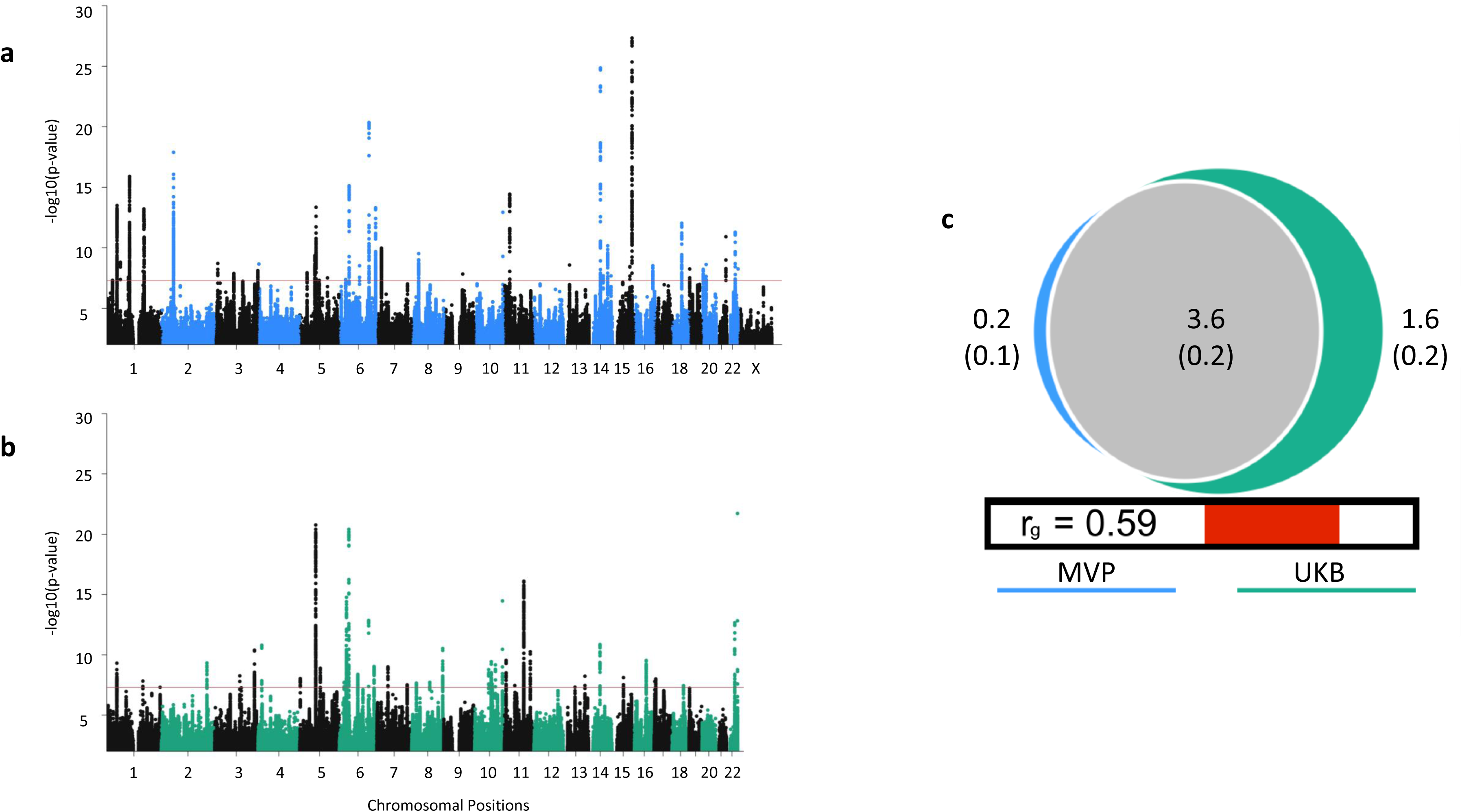
Comparison of SNHL between the clinically diagnosed MVP and self-reported UKB GWAS. **a-b)** Manhattan plots of sensorineural hearing loss (SNHL) GWAS. The red line represents genome-wide significance (GWS) at p < 5 × 10−8. a) MVP GWAS of clinically diagnosed SNHL in European ancestry participants (176,393 cases and 172,117 controls) identified 43 GWS loci. b) UKB GWAS of self-reported SNHL (87,056 cases and 163,333 controls) identified 41 GWS loci. The X chromosome was not analyzed in the UKB GWAS. **c)** Quantification of the polygenic overlap between SNHL in the MVP and UKB. Bivariate MiXeR modeling estimated 3,624 causal variants are shared between cohorts (gray). There was only a small amount of polygenicity unique to MVP (blue), such that shared polygenicity accounted for 95.6% of the causal variants influencing MVP. The UKB had a larger unique polygenic component (green), such that shared polygenicity accounted for 69.2% of the causal variants influencing UKB. The numbers (in thousands, with standard errors in parenthesis) in the Venn diagram indicate the estimated quantity of causal variants per component. The size of the circles reflects the degree of polygenicity. Genetic correlation (rg) estimated between the two phenotypes is shown below the Venn diagram.

### Gene-based, gene-set, and gene-tissue analyses in MVP

To aggregate genetic markers at the level of genes and allow integration with gene expression data, we performed gene-based analyses using MAGMA^42^. Gene-based analyses identified a total of 76 GWS genes across the EUA, AFA, IAA, and meta-analyses (Supplementary Table 4, Fig. S2 and S3a), several of which had not already been implicated by the positional mapping of SNPs to genes. To ascertain specific biological pathways, a competitive set-based analysis of 17,012 pre-defined curated gene sets and gene ontology (GO) terms obtained from MsigDB was performed in the MVP meta-analysis (Supplementary Table 5). Significant curated gene sets identified were in the gene ontology biological processes area, i.e., GOBP_EAR_DEVELOPMENT (p = 1.52E-07), GOBP_SENSORY_PERCEPTION_OF_ MECHANICAL_STIMULUS (p = 3.19E-07), and NIKOLSKY_BREAST_CANCER_14Q22_AMPLICON (p = 2.50E-06). We note that the Nikolsky breast cancer amplicon pathway significance level was primarily driven by one locus with two mapped genes with SNPs in partial LD, *NID2* (p-value 2.61E-14) and *C14orf116* (*FOXN3,* p = 7.25E-10) (Supplementary Table 6).

### Genetic comparisons of MVP and UKB SNHL GWAS

We performed a comparison of the genetic architecture of SNHL based on the GWAS using ICD codes in MVP with a similarly sized GWAS (cases: N = 87,056; controls: N = 163,333) based on self-report in the previously published UKB GWAS^12^, which had identified 41 GWS loci (Fig. 1b). Only data of EUA individuals were included.

The *h^2^_SNP_* estimated from the MVP was 0.0761 (SE = 0.0039, on the liability scale at 30% population prevalence) (Supplementary Table 7A). In contrast, *h^2^_SNP_* estimated from the UKB was higher at 0.1162 (SE = 0.0059, liability scale at 30% population prevalence). Differences in genetic architecture were further explored with univariate MiXeR, noting that *h^2^_SNP_*is proportional to the number of causal variants and the effects sizes of these variants. Univariate MiXeR analyses identified greater polygenicity within UKB (5,240 causal variants, SE = 250) than in MVP (3,790 causal variants, SE = 198) (*z* = 4.5, p = 5.4 x 10^-6^). However, variant effect sizes were not significantly different between studies (*z*-test comparing causal effect size variance estimates: *z* = 1.3, p = 0.18) (Fig. 1c and Supplementary Table 8a).

Shared and unshared genetic variation between studies was explored with bivariate MiXeR analyses. MiXeR estimated that MVP and UKB shared 3,624 (SE = 204) causal variants, comprising 95.6% of the total MVP causal variants, but only 69.2% of the total UKB causal variants (Fig. 1c and Supplementary Table 8b). Correspondingly, MiXeR estimated that 1,617 (SE = 235) variants were unique to the UKB, but only 167 (SE = 126) variants were unique to MVP.

Genetic correlation among shared variants was r_g_ = 0.73 (SE = 0.03), which was higher than the standard genome-wide estimate of genetic correlation r_g_ = 0.59 (SE = 0.03) estimated by LDSC (Supplementary Table 7B). A comparison of only GWS loci showed a correlation of *r* = 0.77 between effect sizes of leading variants, further establishing that the identified GWS loci have similar effects across studies. In conclusion, while self-reported hearing loss in the UKB is influenced by a broader set of variation compared to the clinically diagnosed SNHL in MVP, there is a substantial core set of variation with comparable effect sizes across these cohorts. Of note however, our analyses are not able to distinguish between differences in populations or the assessment tools used.

### Meta-analysis of MVP and UKB EUA GWAS

An inverse variance weighted fixed effects meta-analysis in 598,899 EUA individuals (cases: N = 263,449; controls: N = 335,450) of the EUA MVP and UKB GWAS identified 99 GWS loci (Supplementary Table 9). Of these, 43 were not previously identified in either the MVP or UKB GWAS. These data will be used in downstream analyses that are based on linkage disequilibrium (LD) patterns, as many of the non-EUA individuals in the MVP are admixed.

### Multi-ancestry meta-analysis of MVP and UKB

A meta-analysis of the MVP multi-ancestry GWAS and the UKB GWAS identified a total of 108 GWS loci in 725,904 individuals; 54 of these loci have a novel protein-coding gene and/or FLAMES prioritized gene (see definition of ’novel’ in methods), while two loci (locus 15 and 49) did not include a gene (Table 1, Supplementary Table 10 and Fig. 2b). Regional association plots comparing ancestry-specific analyses for these loci are shown in Supplementary Data 1. Functional mapping and annotation of risk loci using the FUMA pipeline are summarized in Supplementary Table 11. Of the 108 loci, 26 (24%) contained an exonic GWS SNP with a predicted deleterious CADD score (>12.37).

**Figure 2.**
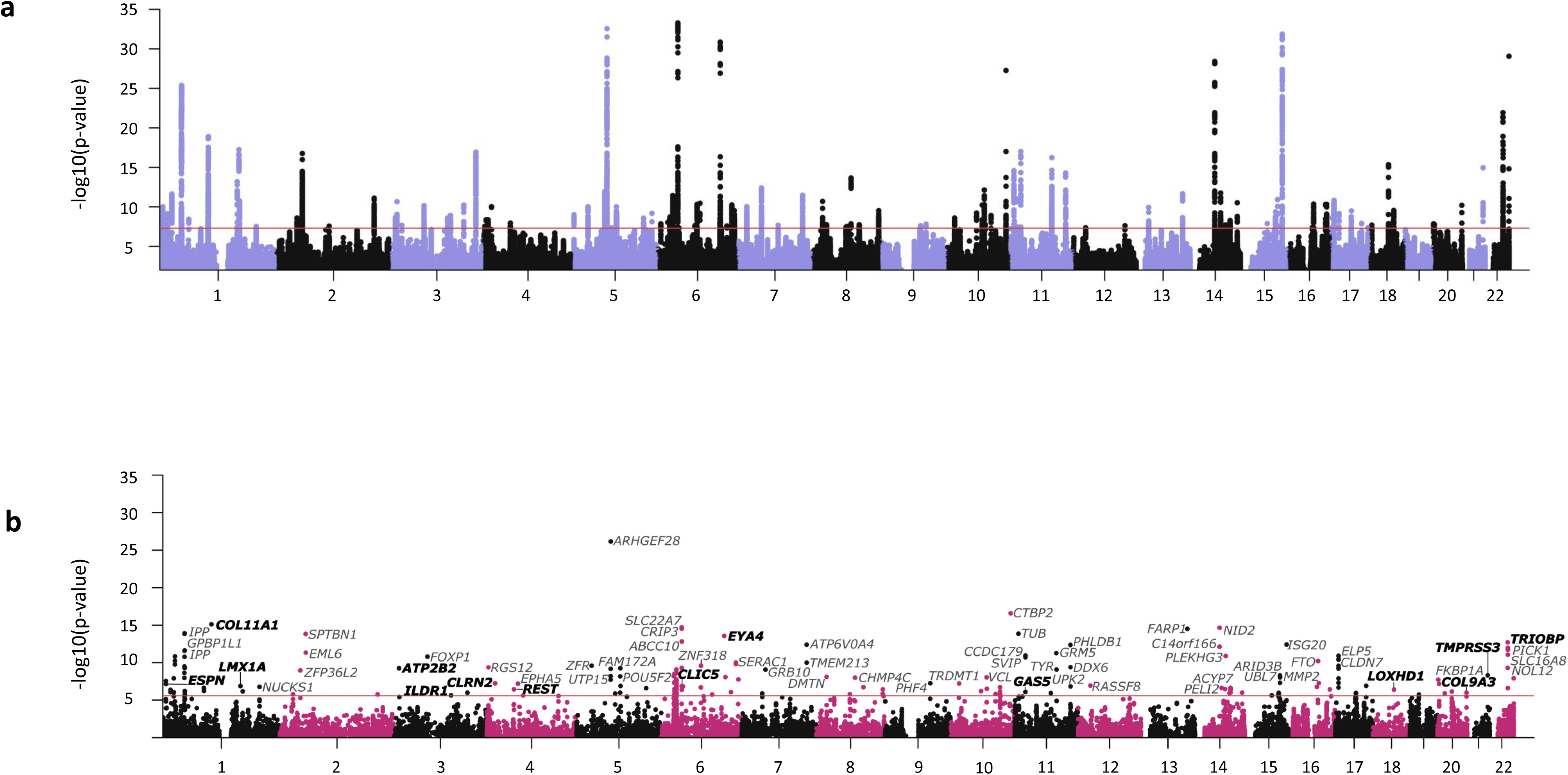
Risk loci (N = 108) and gene discovery (N = 192) for SNHL in a multi-ancestry meta-analysis. Manhattan plots of SNP **a)** and gene-level **b)** associations from meta-analysis of the MVP multi-ancestry and UKB sensorineural hearing loss (SNHL) GWAS (297,296 cases and 428,608 controls). The red line represents genome-wide significance at p < 5 × 10−8 (panel a) and gene-wide significance p < 2.66 × 10−6 (panel b, Bonferroni correction for 18,809 genes tested), respectively. A selection of GWS genes are labeled, and genes listed in HereditaryHearingLoss.org are labeled in bold font.

**Figure 3.**
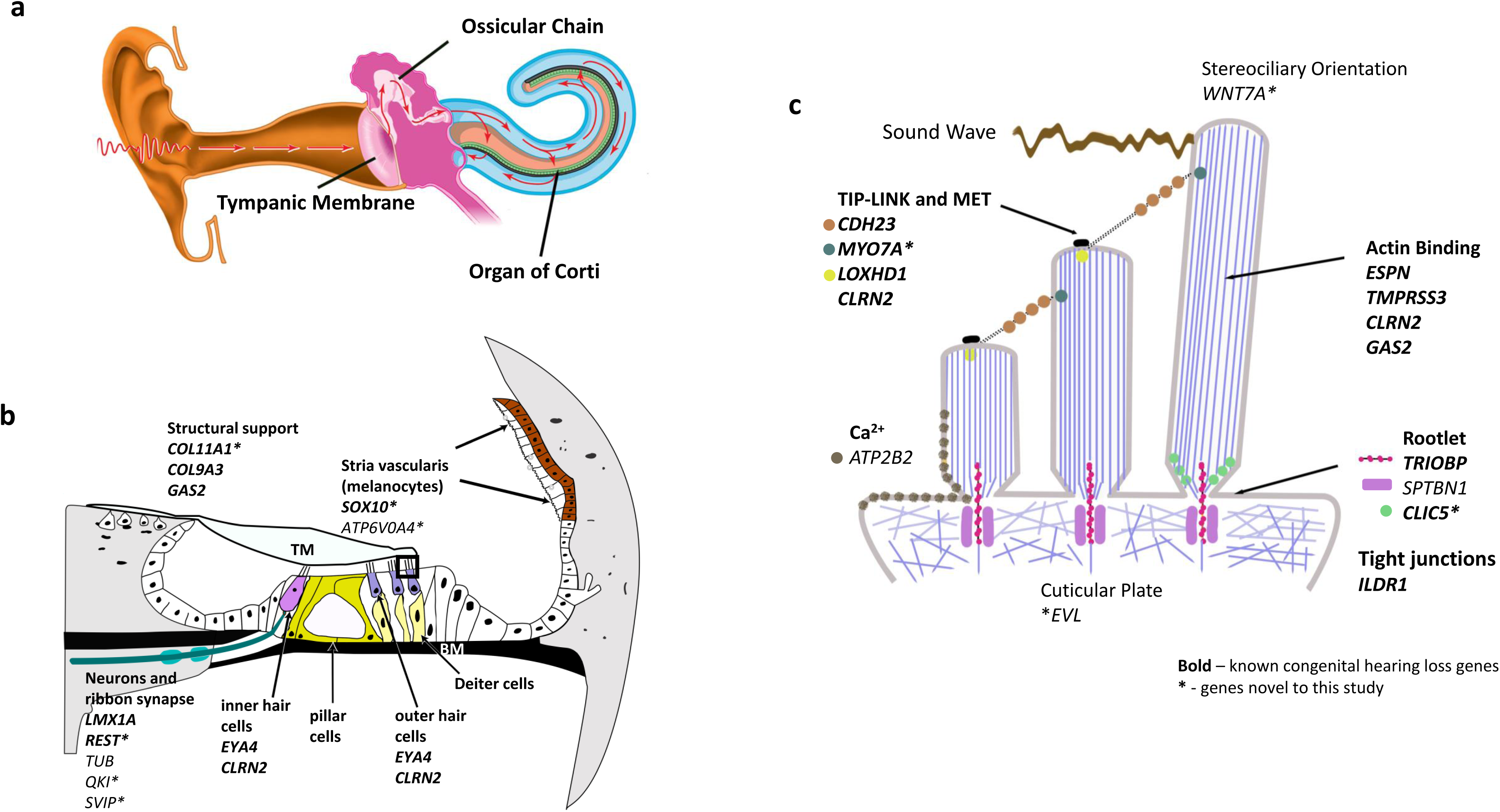
Putative influences of significant hearing loss genes on cochlear function. **a)** Cross-section of ear. Sound waves travel through the external auditory canal, vibrating the tympanic membrane and ossicular chain, continuing as a fluid wave into the cochlea. The Organ of Corti (OC) is suspended within the center of the cochlea (green). **b)** Cross-section of the OC. supported by the basilar membrane below (BM). Movement is restricted by the tectorial membrane above. The shearing motion of the fluid endolymph wave motion against OHC stereocilia induces a dampening, focusing the wave towards the locally juxtaposed inner hair cells (IHCs). IHCs then transduce this movement into an electric signal, releasing neurotransmitter at the ribbon synapse. Post-synaptic neurons of the cochlear nerve transmit signals to the auditory cortex. The stria vascularis maintains the electric potential of endolymph that bathes the apices of hair cells. **c)** Expanded area of apex of hair cells from b., showing stereociliary movement of both OHCs and IHCs. Fluid/sound wave movement pushes stereocilia laterally, stretching tip-links attaching the taller stereocilium to its adjacent shorter one. The stereocilium’s parallel f-actin fibers are critical for stiffness and rigidity of the stereocilia, while allowing for bending at the reinforced rootlet at the attachment of the stereocilia to the apical surface of the hair cell. The rootlet is the focus area of vibration secondary to sound as measured by high-resolution optics^73^. Tip-links consist of chained protein threads connecting sequential rows of stereociliae, from the top of the shorter stereocilia to the side of the next taller structure This movement opens a mechanoelectrical transduction (MET) channel at its tip (black), allowing influx of Ca++ and K+ and triggering depolarization at the base of the cell. Tight junctions between hair cells and supporting cells separate endolymph bathing the stereocilia from its cell bodies. Genes that are not color-coded are ubiquitous, involved in development and/or maintenance.

**Table 1.**
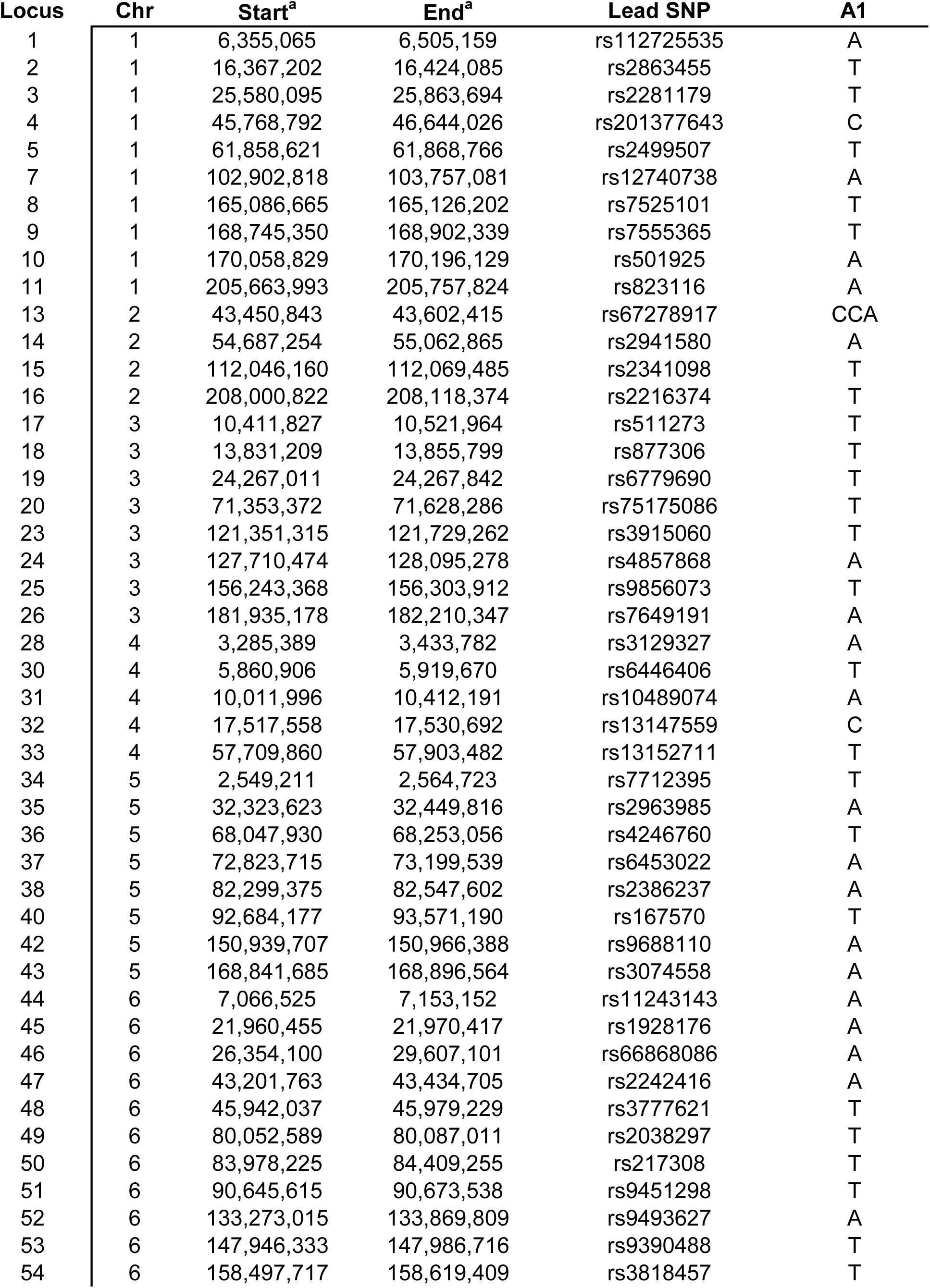
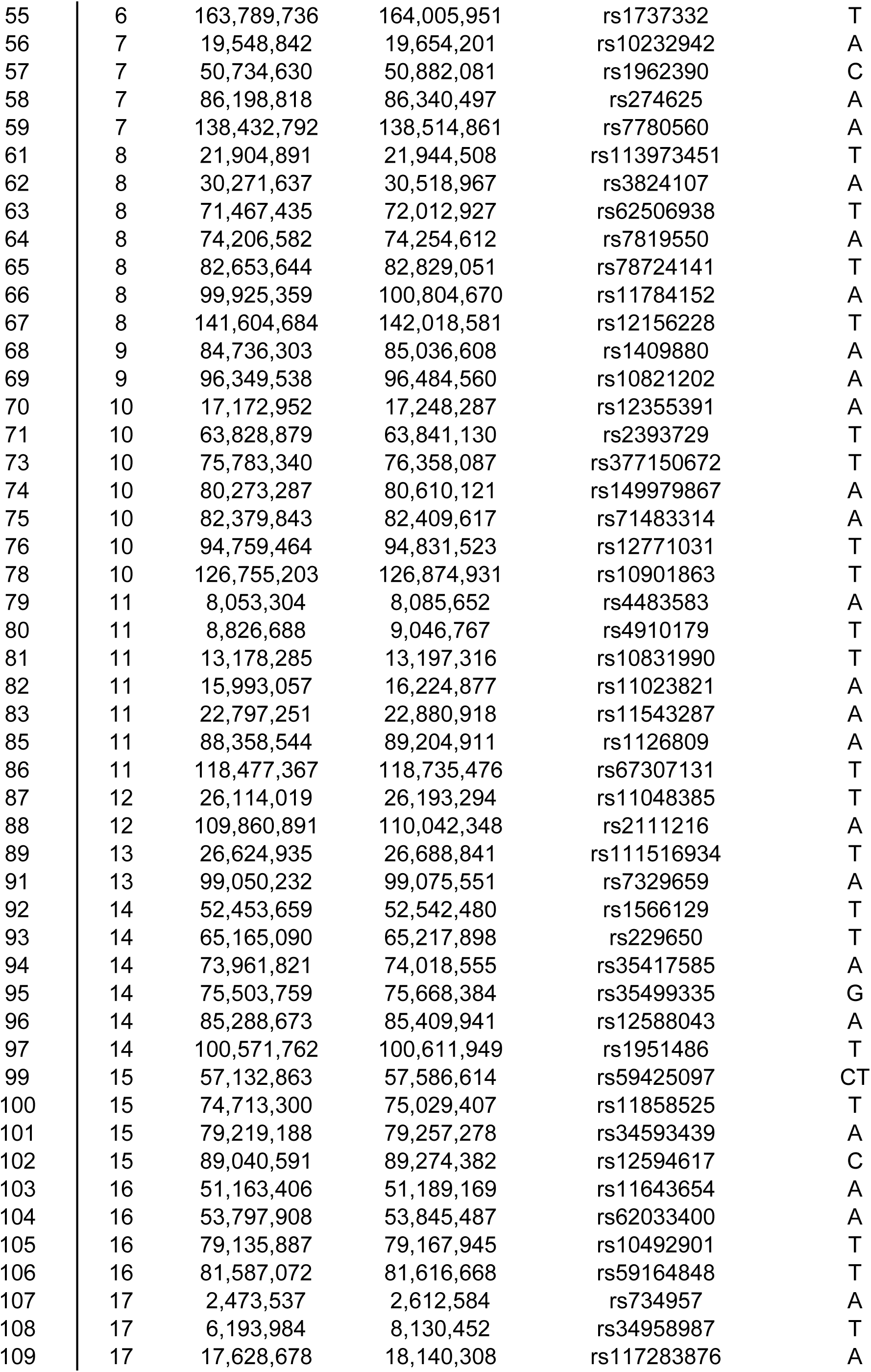
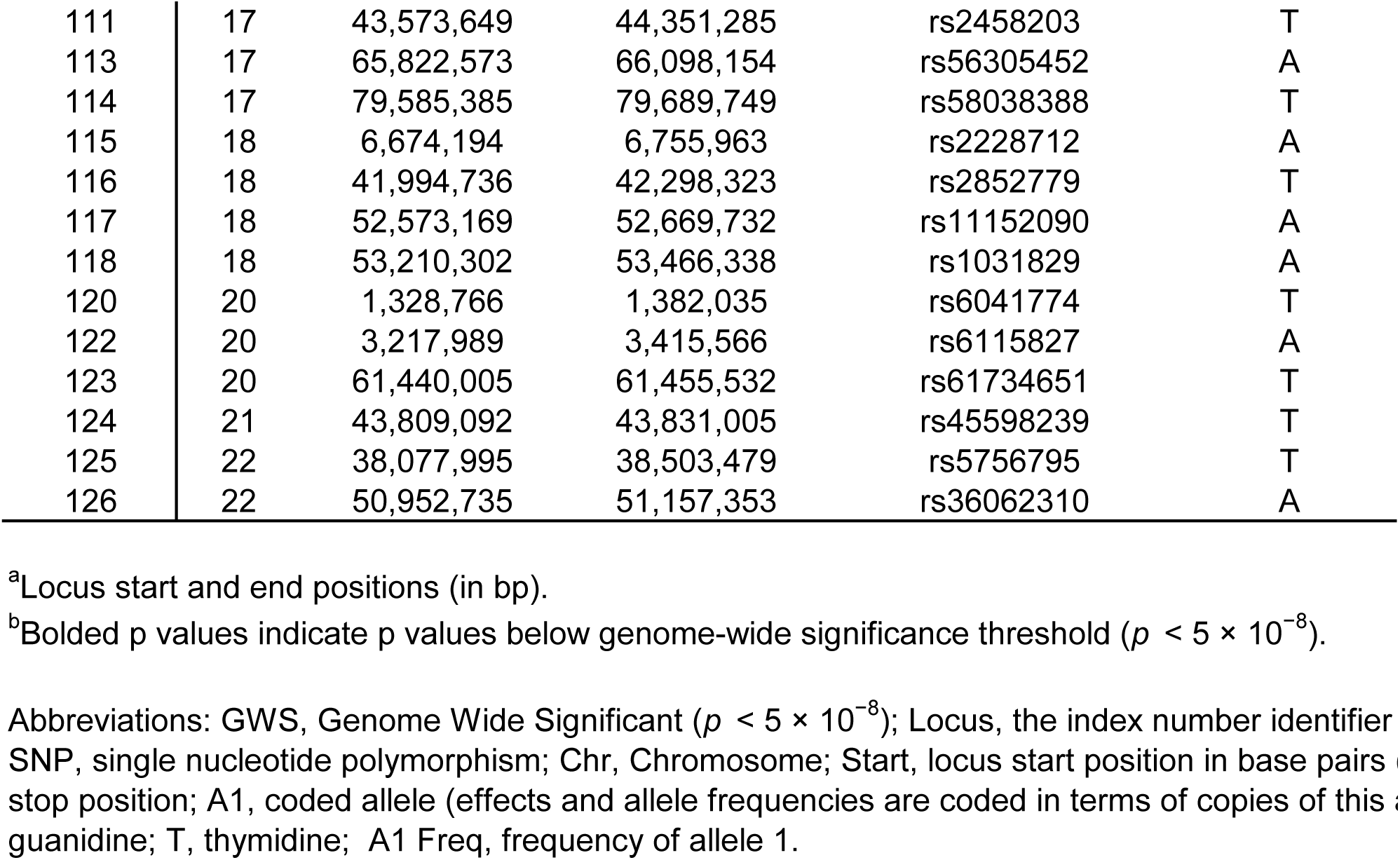
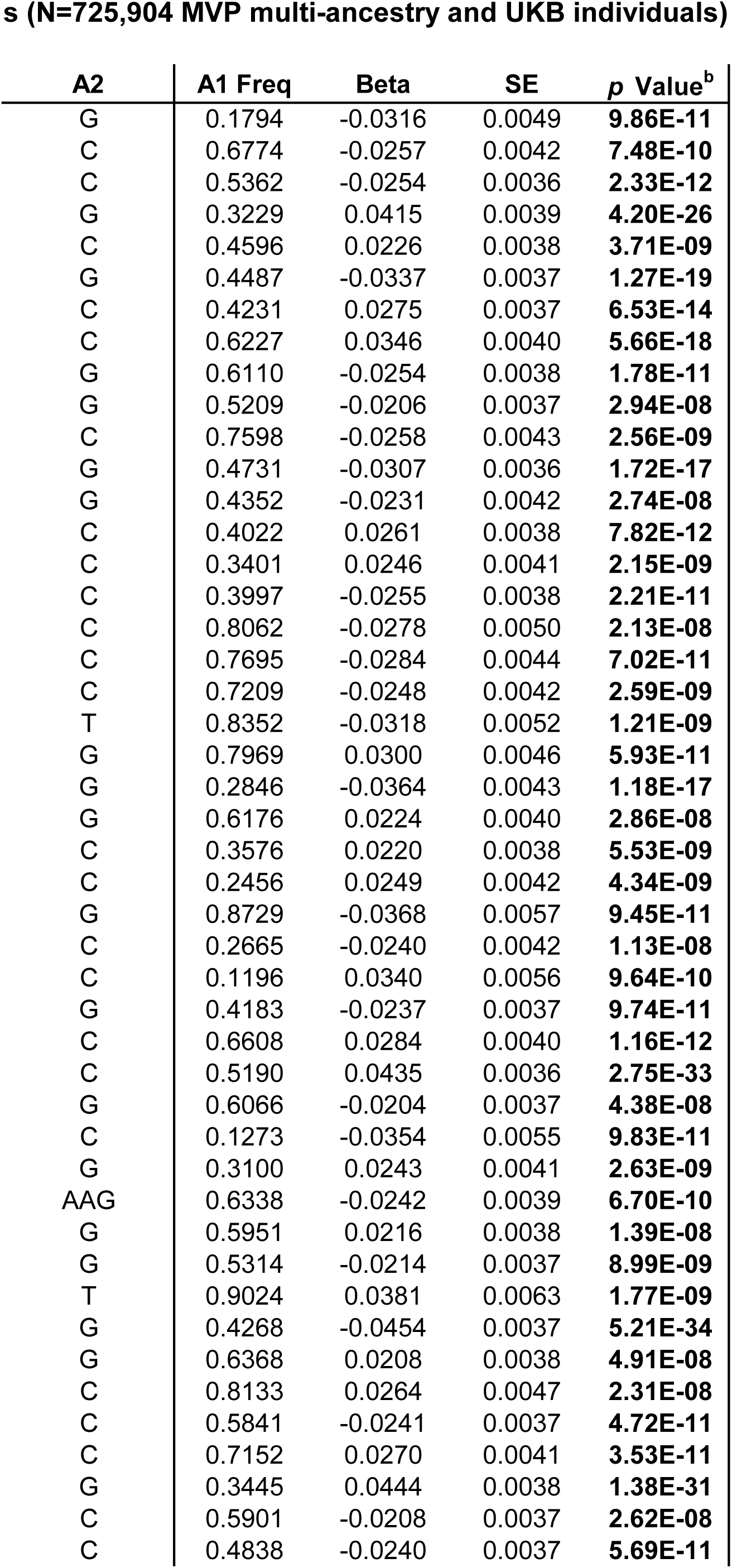
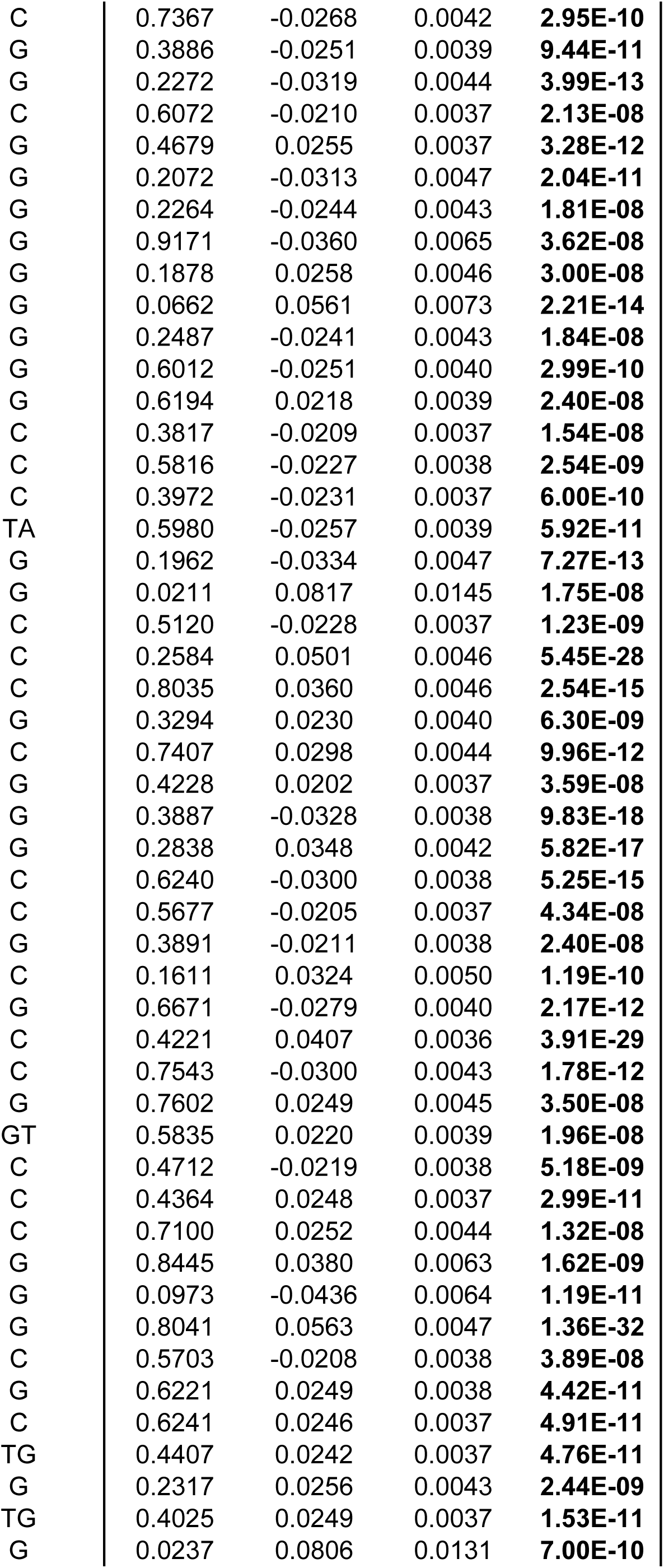
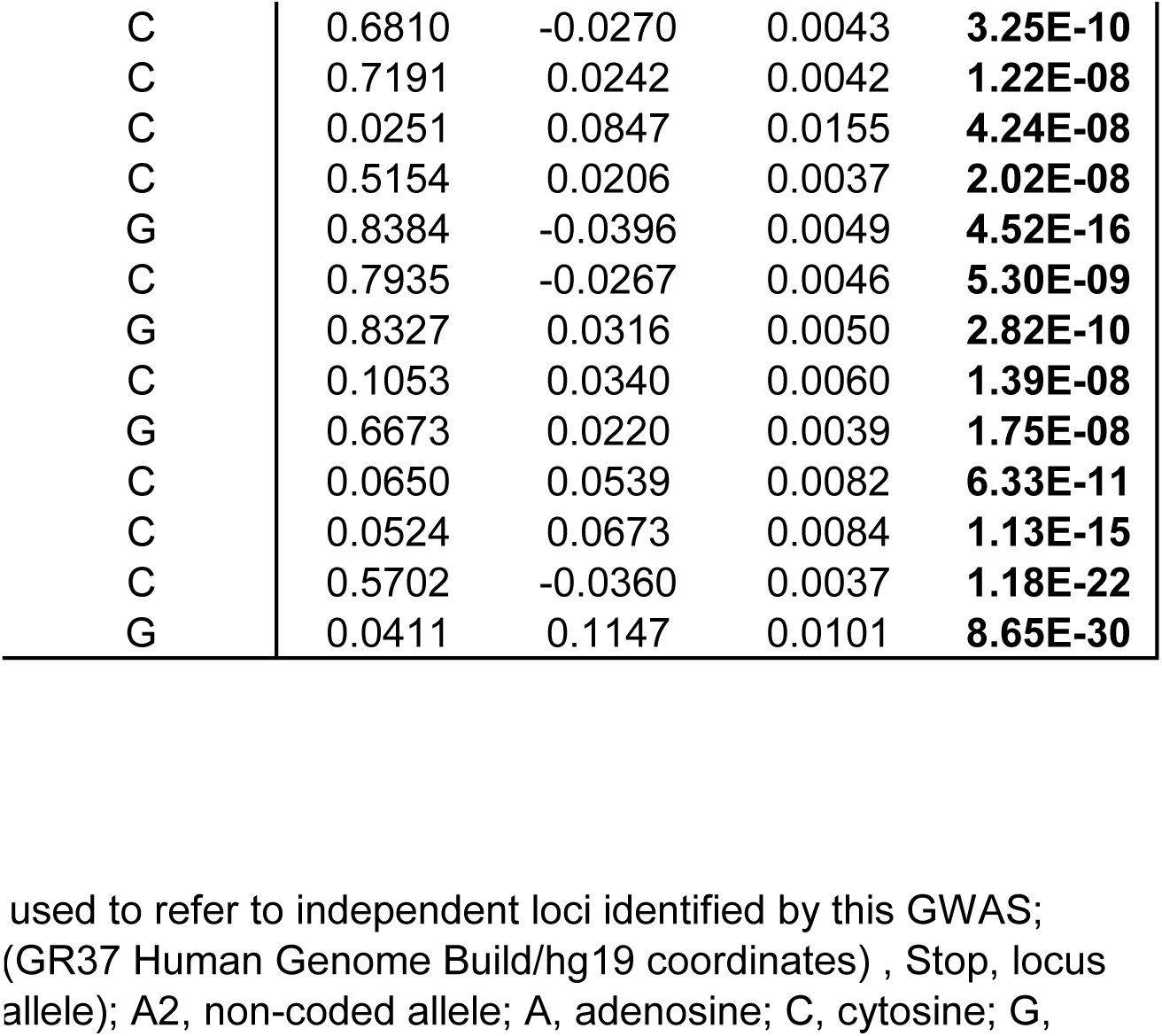
Genome-wide significant loci in meta-analysis of sensorineural hearing los.

| **Locus** | **Chr** | **Start<sup>a</sup>** | **End<sup>a</sup>** | **Lead SNP** | **A1** |
| --- | --- | --- | --- | --- | --- |
| 1 | 1 | 6,355,065 | 6,505,159 | rs112725535 | A |
| 2 | 1 | 16,367,202 | 16,424,085 | rs2863455 | T |
| 3 | 1 | 25,580,095 | 25,863,694 | rs2281179 | T |
| 4 | 1 | 45,768,792 | 46,644,026 | rs201377643 | C |
| 5 | 1 | 61,858,621 | 61,868,766 | rs2499507 | T |
| 7 | 1 | 102,902,818 | 103,757,081 | rs12740738 | A |
| 8 | 1 | 165,086,665 | 165,126,202 | rs7525101 | T |
| 9 | 1 | 168,745,350 | 168,902,339 | rs7555365 | T |
| 10 | 1 | 170,058,829 | 170,196,129 | rs501925 | A |
| 11 | 1 | 205,663,993 | 205,757,824 | rs823116 | A |
| 13 | 2 | 43,450,843 | 43,602,415 | rs67278917 | CCA |
| 14 | 2 | 54,687,254 | 55,062,865 | rs2941580 | A |
| 15 | 2 | 112,046,160 | 112,069,485 | rs2341098 | T |
| 16 | 2 | 208,000,822 | 208,118,374 | rs2216374 | T |
| 17 | 3 | 10,411,827 | 10,521,964 | rs511273 | T |
| 18 | 3 | 13,831,209 | 13,855,799 | rs877306 | T |
| 19 | 3 | 24,267,011 | 24,267,842 | rs6779690 | T |
| 20 | 3 | 71,353,372 | 71,628,286 | rs75175086 | T |
| 23 | 3 | 121,351,315 | 121,729,262 | rs3915060 | T |
| 24 | 3 | 127,710,474 | 128,095,278 | rs4857868 | A |
| 25 | 3 | 156,243,368 | 156,303,912 | rs9856073 | T |
| 26 | 3 | 181,935,178 | 182,210,347 | rs7649191 | A |
| 28 | 4 | 3,285,389 | 3,433,782 | rs3129327 | A |
| 30 | 4 | 5,860,906 | 5,919,670 | rs6446406 | T |
| 31 | 4 | 10,011,996 | 10,412,191 | rs10489074 | A |
| 32 | 4 | 17,517,558 | 17,530,692 | rs13147559 | C |
| 33 | 4 | 57,709,860 | 57,903,482 | rs13152711 | T |
| 34 | 5 | 2,549,211 | 2,564,723 | rs7712395 | T |
| 35 | 5 | 32,323,623 | 32,449,816 | rs2963985 | A |
| 36 | 5 | 68,047,930 | 68,253,056 | rs4246760 | T |
| 37 | 5 | 72,823,715 | 73,199,539 | rs6453022 | A |
| 38 | 5 | 82,299,375 | 82,547,602 | rs2386237 | A |
| 40 | 5 | 92,684,177 | 93,571,190 | rs167570 | T |
| 42 | 5 | 150,939,707 | 150,966,388 | rs9688110 | A |
| 43 | 5 | 168,841,685 | 168,896,564 | rs3074558 | A |
| 44 | 6 | 7,066,525 | 7,153,152 | rs11243143 | A |
| 45 | 6 | 21,960,455 | 21,970,417 | rs1928176 | A |
| 46 | 6 | 26,354,100 | 29,607,101 | rs66868086 | A |
| 47 | 6 | 43,201,763 | 43,434,705 | rs2242416 | A |
| 48 | 6 | 45,942,037 | 45,979,229 | rs3777621 | T |
| 49 | 6 | 80,052,589 | 80,087,011 | rs2038297 | T |
| 50 | 6 | 83,978,225 | 84,409,255 | rs217308 | T |
| 51 | 6 | 90,645,615 | 90,673,538 | rs9451298 | T |
| 52 | 6 | 133,273,015 | 133,869,809 | rs9493627 | A |
| 53 | 6 | 147,946,333 | 147,986,716 | rs9390488 | T |
| 54 | 6 | 158,497,717 | 158,619,409 | rs3818457 | T |
| 55 | 6 | 163,789,736 | 164,005,951 | rs1737332 | T |
| 56 | 7 | 19,548,842 | 19,654,201 | rs10232942 | A |
| 57 | 7 | 50,734,630 | 50,882,081 | rs1962390 | C |
| 58 | 7 | 86,198,818 | 86,340,497 | rs274625 | A |
| 59 | 7 | 138,432,792 | 138,514,861 | rs7780560 | A |
| 61 | 8 | 21,904,891 | 21,944,508 | rs113973451 | T |
| 62 | 8 | 30,271,637 | 30,518,967 | rs3824107 | A |
| 63 | 8 | 71,467,435 | 72,012,927 | rs62506938 | T |
| 64 | 8 | 74,206,582 | 74,254,612 | rs7819550 | A |
| 65 | 8 | 82,653,644 | 82,829,051 | rs78724141 | T |
| 66 | 8 | 99,925,359 | 100,804,670 | rs11784152 | A |
| 67 | 8 | 141,604,684 | 142,018,581 | rs12156228 | T |
| 68 | 9 | 84,736,303 | 85,036,608 | rs1409880 | A |
| 69 | 9 | 96,349,538 | 96,484,560 | rs10821202 | A |
| 70 | 10 | 17,172,952 | 17,248,287 | rs12355391 | A |
| 71 | 10 | 63,828,879 | 63,841,130 | rs2393729 | T |
| 73 | 10 | 75,783,340 | 76,358,087 | rs377150672 | T |
| 74 | 10 | 80,273,287 | 80,610,121 | rs149979867 | A |
| 75 | 10 | 82,379,843 | 82,409,617 | rs71483314 | A |
| 76 | 10 | 94,759,464 | 94,831,523 | rs12771031 | T |
| 78 | 10 | 126,755,203 | 126,874,931 | rs10901863 | T |
| 79 | 11 | 8,053,304 | 8,085,652 | rs4483583 | A |
| 80 | 11 | 8,826,688 | 9,046,767 | rs4910179 | T |
| 81 | 11 | 13,178,285 | 13,197,316 | rs10831990 | T |
| 82 | 11 | 15,993,057 | 16,224,877 | rs11023821 | A |
| 83 | 11 | 22,797,251 | 22,880,918 | rs11543287 | A |
| 85 | 11 | 88,358,544 | 89,204,911 | rs1126809 | A |
| 86 | 11 | 118,477,367 | 118,735,476 | rs67307131 | T |
| 87 | 12 | 26,114,019 | 26,193,294 | rs11048385 | T |
| 88 | 12 | 109,860,891 | 110,042,348 | rs2111216 | A |
| 89 | 13 | 26,624,935 | 26,688,841 | rs111516934 | T |
| 91 | 13 | 99,050,232 | 99,075,551 | rs7329659 | A |
| 92 | 14 | 52,453,659 | 52,542,480 | rs1566129 | T |
| 93 | 14 | 65,165,090 | 65,217,898 | rs229650 | T |
| 94 | 14 | 73,961,821 | 74,018,555 | rs35417585 | A |
| 95 | 14 | 75,503,759 | 75,668,384 | rs35499335 | G |
| 96 | 14 | 85,288,673 | 85,409,941 | rs12588043 | A |
| 97 | 14 | 100,571,762 | 100,611,949 | rs1951486 | T |
| 99 | 15 | 57,132,863 | 57,586,614 | rs59425097 | CT |
| 100 | 15 | 74,713,300 | 75,029,407 | rs11858525 | T |
| 101 | 15 | 79,219,188 | 79,257,278 | rs34593439 | A |
| 102 | 15 | 89,040,591 | 89,274,382 | rs12594617 | C |
| 103 | 16 | 51,163,406 | 51,189,169 | rs11643654 | A |
| 104 | 16 | 53,797,908 | 53,845,487 | rs62033400 | A |
| 105 | 16 | 79,135,887 | 79,167,945 | rs10492901 | T |
| 106 | 16 | 81,587,072 | 81,616,668 | rs59164848 | T |
| 107 | 17 | 2,473,537 | 2,612,584 | rs734957 | A |
| 108 | 17 | 6,193,984 | 8,130,452 | rs34958987 | T |
| 109 | 17 | 17,628,678 | 18,140,308 | rs117283876 | A |
| 111 | 17 | 43,573,649 | 44,351,285 | rs2458203 | T |
| 113 | 17 | 65,822,573 | 66,098,154 | rs56305452 | A |
| 114 | 17 | 79,585,385 | 79,689,749 | rs58038388 | T |
| 115 | 18 | 6,674,194 | 6,755,963 | rs2228712 | A |
| 116 | 18 | 41,994,736 | 42,298,323 | rs2852779 | T |
| 117 | 18 | 52,573,169 | 52,669,732 | rs11152090 | A |
| 118 | 18 | 53,210,302 | 53,466,338 | rs1031829 | A |
| 120 | 20 | 1,328,766 | 1,382,035 | rs6041774 | T |
| 122 | 20 | 3,217,989 | 3,415,566 | rs6115827 | A |
| 123 | 20 | 61,440,005 | 61,455,532 | rs61734651 | T |
| 124 | 21 | 43,809,092 | 43,831,005 | rs45598239 | T |
| 125 | 22 | 38,077,995 | 38,503,479 | rs5756795 | T |
| 126 | 22 | 50,952,735 | 51,157,353 | rs36062310 | A |

<sup>a</sup>Locus start and end positions (in bp).
<sup>b</sup>Bolded p values indicate p values below genome-wide significance threshold ( $p < 5 \times 10^{-8}$ ).
Abbreviations: GWS, Genome Wide Significant ( $p < 5 \times 10^{-8}$ ); Locus, the index number identifier; SNP, single nucleotide polymorphism; Chr, Chromosome; Start, locus start position in base pairs; Stop, locus stop position; A1, coded allele (effects and allele frequencies are coded in terms of copies of this allele); G, guanine; A, adenine; C, cytosine; T, thymidine; A1 Freq, frequency of allele 1.

**s (N=725,904 MVP multi-ancestry and UKB individuals)**
| **A2** | **A1 Freq** | **Beta** | **SE** | **p Value<sup>b</sup>** |
| --- | --- | --- | --- | --- |
| G | 0.1794 | -0.0316 | 0.0049 | **9.86E-11** |
| C | 0.6774 | -0.0257 | 0.0042 | **7.48E-10** |
| C | 0.5362 | -0.0254 | 0.0036 | **2.33E-12** |
| G | 0.3229 | 0.0415 | 0.0039 | **4.20E-26** |
| C | 0.4596 | 0.0226 | 0.0038 | **3.71E-09** |
| G | 0.4487 | -0.0337 | 0.0037 | **1.27E-19** |
| C | 0.4231 | 0.0275 | 0.0037 | **6.53E-14** |
| C | 0.6227 | 0.0346 | 0.0040 | **5.66E-18** |
| G | 0.6110 | -0.0254 | 0.0038 | **1.78E-11** |
| G | 0.5209 | -0.0206 | 0.0037 | **2.94E-08** |
| C | 0.7598 | -0.0258 | 0.0043 | **2.56E-09** |
| G | 0.4731 | -0.0307 | 0.0036 | **1.72E-17** |
| G | 0.4352 | -0.0231 | 0.0042 | **2.74E-08** |
| C | 0.4022 | 0.0261 | 0.0038 | **7.82E-12** |
| C | 0.3401 | 0.0246 | 0.0041 | **2.15E-09** |
| C | 0.3997 | -0.0255 | 0.0038 | **2.21E-11** |
| C | 0.8062 | -0.0278 | 0.0050 | **2.13E-08** |
| C | 0.7695 | -0.0284 | 0.0044 | **7.02E-11** |
| C | 0.7209 | -0.0248 | 0.0042 | **2.59E-09** |
| T | 0.8352 | -0.0318 | 0.0052 | **1.21E-09** |
| G | 0.7969 | 0.0300 | 0.0046 | **5.93E-11** |
| G | 0.2846 | -0.0364 | 0.0043 | **1.18E-17** |
| G | 0.6176 | 0.0224 | 0.0040 | **2.86E-08** |
| C | 0.3576 | 0.0220 | 0.0038 | **5.53E-09** |
| C | 0.2456 | 0.0249 | 0.0042 | **4.34E-09** |
| G | 0.8729 | -0.0368 | 0.0057 | **9.45E-11** |
| C | 0.2665 | -0.0240 | 0.0042 | **1.13E-08** |
| C | 0.1196 | 0.0340 | 0.0056 | **9.64E-10** |
| G | 0.4183 | -0.0237 | 0.0037 | **9.74E-11** |
| C | 0.6608 | 0.0284 | 0.0040 | **1.16E-12** |
| C | 0.5190 | 0.0435 | 0.0036 | **2.75E-33** |
| G | 0.6066 | -0.0204 | 0.0037 | **4.38E-08** |
| C | 0.1273 | -0.0354 | 0.0055 | **9.83E-11** |
| G | 0.3100 | 0.0243 | 0.0041 | **2.63E-09** |
| AAG | 0.6338 | -0.0242 | 0.0039 | **6.70E-10** |
| G | 0.5951 | 0.0216 | 0.0038 | **1.39E-08** |
| G | 0.5314 | -0.0214 | 0.0037 | **8.99E-09** |
| T | 0.9024 | 0.0381 | 0.0063 | **1.77E-09** |
| G | 0.4268 | -0.0454 | 0.0037 | **5.21E-34** |
| G | 0.6368 | 0.0208 | 0.0038 | **4.91E-08** |
| C | 0.8133 | 0.0264 | 0.0047 | **2.31E-08** |
| C | 0.5841 | -0.0241 | 0.0037 | **4.72E-11** |
| C | 0.7152 | 0.0270 | 0.0041 | **3.53E-11** |
| G | 0.3445 | 0.0444 | 0.0038 | **1.38E-31** |
| C | 0.5901 | -0.0208 | 0.0037 | **2.62E-08** |
| C | 0.4838 | -0.0240 | 0.0037 | **5.69E-11** |
| C | 0.7367 | -0.0268 | 0.0042 | **2.95E-10** |
| G | 0.3886 | -0.0251 | 0.0039 | **9.44E-11** |
| G | 0.2272 | -0.0319 | 0.0044 | **3.99E-13** |
| C | 0.6072 | -0.0210 | 0.0037 | **2.13E-08** |
| G | 0.4679 | 0.0255 | 0.0037 | **3.28E-12** |
| G | 0.2072 | -0.0313 | 0.0047 | **2.04E-11** |
| G | 0.2264 | -0.0244 | 0.0043 | **1.81E-08** |
| G | 0.9171 | -0.0360 | 0.0065 | **3.62E-08** |
| G | 0.1878 | 0.0258 | 0.0046 | **3.00E-08** |
| G | 0.0662 | 0.0561 | 0.0073 | **2.21E-14** |
| G | 0.2487 | -0.0241 | 0.0043 | **1.84E-08** |
| G | 0.6012 | -0.0251 | 0.0040 | **2.99E-10** |
| G | 0.6194 | 0.0218 | 0.0039 | **2.40E-08** |
| C | 0.3817 | -0.0209 | 0.0037 | **1.54E-08** |
| C | 0.5816 | -0.0227 | 0.0038 | **2.54E-09** |
| C | 0.3972 | -0.0231 | 0.0037 | **6.00E-10** |
| TA | 0.5980 | -0.0257 | 0.0039 | **5.92E-11** |
| G | 0.1962 | -0.0334 | 0.0047 | **7.27E-13** |
| G | 0.0211 | 0.0817 | 0.0145 | **1.75E-08** |
| C | 0.5120 | -0.0228 | 0.0037 | **1.23E-09** |
| C | 0.2584 | 0.0501 | 0.0046 | **5.45E-28** |
| C | 0.8035 | 0.0360 | 0.0046 | **2.54E-15** |
| G | 0.3294 | 0.0230 | 0.0040 | **6.30E-09** |
| C | 0.7407 | 0.0298 | 0.0044 | **9.96E-12** |
| G | 0.4228 | 0.0202 | 0.0037 | **3.59E-08** |
| G | 0.3887 | -0.0328 | 0.0038 | **9.83E-18** |
| G | 0.2838 | 0.0348 | 0.0042 | **5.82E-17** |
| C | 0.6240 | -0.0300 | 0.0038 | **5.25E-15** |
| C | 0.5677 | -0.0205 | 0.0037 | **4.34E-08** |
| G | 0.3891 | -0.0211 | 0.0038 | **2.40E-08** |
| C | 0.1611 | 0.0324 | 0.0050 | **1.19E-10** |
| G | 0.6671 | -0.0279 | 0.0040 | **2.17E-12** |
| C | 0.4221 | 0.0407 | 0.0036 | **3.91E-29** |
| C | 0.7543 | -0.0300 | 0.0043 | **1.78E-12** |
| G | 0.7602 | 0.0249 | 0.0045 | **3.50E-08** |
| GT | 0.5835 | 0.0220 | 0.0039 | **1.96E-08** |
| C | 0.4712 | -0.0219 | 0.0038 | **5.18E-09** |
| C | 0.4364 | 0.0248 | 0.0037 | **2.99E-11** |
| C | 0.7100 | 0.0252 | 0.0044 | **1.32E-08** |
| G | 0.8445 | 0.0380 | 0.0063 | **1.62E-09** |
| G | 0.0973 | -0.0436 | 0.0064 | **1.19E-11** |
| G | 0.8041 | 0.0563 | 0.0047 | **1.36E-32** |
| C | 0.5703 | -0.0208 | 0.0038 | **3.89E-08** |
| G | 0.6221 | 0.0249 | 0.0038 | **4.42E-11** |
| C | 0.6241 | 0.0246 | 0.0037 | **4.91E-11** |
| TG | 0.4407 | 0.0242 | 0.0037 | **4.76E-11** |
| G | 0.2317 | 0.0256 | 0.0043 | **2.44E-09** |
| TG | 0.4025 | 0.0249 | 0.0037 | **1.53E-11** |
| G | 0.0237 | 0.0806 | 0.0131 | **7.00E-10** |
| C | 0.6810 | -0.0270 | 0.0043 | **3.25E-10** |
| C | 0.7191 | 0.0242 | 0.0042 | **1.22E-08** |
| C | 0.0251 | 0.0847 | 0.0155 | **4.24E-08** |
| C | 0.5154 | 0.0206 | 0.0037 | **2.02E-08** |
| G | 0.8384 | -0.0396 | 0.0049 | **4.52E-16** |
| C | 0.7935 | -0.0267 | 0.0046 | **5.30E-09** |
| G | 0.8327 | 0.0316 | 0.0050 | **2.82E-10** |
| C | 0.1053 | 0.0340 | 0.0060 | **1.39E-08** |
| G | 0.6673 | 0.0220 | 0.0039 | **1.75E-08** |
| C | 0.0650 | 0.0539 | 0.0082 | **6.33E-11** |
| C | 0.0524 | 0.0673 | 0.0084 | **1.13E-15** |
| C | 0.5702 | -0.0360 | 0.0037 | **1.18E-22** |
| G | 0.0411 | 0.1147 | 0.0101 | **8.65E-30** |

used to refer to independent loci identified by this GWAS; (GR37 Human Genome Build/hg19 coordinates) , Stop, locus allele); A2, non-coded allele; A, adenosine; C, cytosine; G,

Gene-based analyses (MAGMA) identified 192 genome-wide significant genes, of which 100 are novel (Supplementary Table 12). Competitive set-based analysis of pre-defined curated gene sets and gene ontology (GO) terms identified three significant gene sets, GOBP_SENSORY_PERCEPTION_ OF_MECHANICAL_STIMULUS (p = 6.42E-09), GOMF_ACTIN_BINDING (p = 2.55E-06) and GOMF_CYTOSKELETAL_PROTEIN_BINDING (p = 1.63E-06) (Supplementary Tables 13 and 14).

To identify tissues and pathways underlying these gene-based associations, we performed a MAGMA tissue expression analysis with 31 general human tissue types, including human inner ear tissue from van der Valk *et al*.^44^, and found significant enrichment in the inner ear (*p* = 2.66×10^-8^), nerve (*p* = 1.50×10^-5^) and pituitary (*p* = 8.66×10^-4^) of EUA MVP, UKB, and in meta-analysis. Analysis of 71 different tissue types identified significant enrichment in the cerebellum (*p* = 2.33×10^-5^), cerebellar hemisphere (*p* = 6.22×10^-5^), and tibial nerve (*p* = 3.65×10^-4^), as well as 13 inner ear cell types (Supplementary Table 15 and Fig. S4).

Please note that these gene-tissue analyses were based on GTEx v8 data sets, which do not include the human cochlea, the tissue most relevant to hearing loss. We enriched with human data of one subject only^44^ which had been labeled “utricle”, however, this analysis identified cochlear cell types as well as vestibular tissue. Because of the paucity of human cochlear data, we performed additional enrichment analyses based on available mouse data from two recent, large studies on the organ of Corti^46^ and cochlear cells^45^ (Supplementary Table 16 and Fig. S5). The murine organ of Corti included outer and inner hair cells, pillar cells, Deiter’s cells, and melanocytes. The genetic signal associated with SNHL from the meta-analysis was significantly enriched (p < 0.01) for expression in all cell types (except for Deiter’s cells), conditional on the average expression of cochlear tissues. The cochlear data included 34 cell types. No cell type was significantly enriched when conditioned on the average expression of all cell types as the baseline.

### Examination of implicated genes in the context of congenital hearing loss

The Hereditary Hearing Loss database was used to compare findings from our analyses to genes identified in the context of congenital hearing loss. Of the 196 protein-coding genes (accessed August 9, 2024), 17 genes were implicated by our GWAS from either positional mapping of GWS loci or gene-based analyses (Supplementary Table 17). Twelve out of the 17 are members of significant gene ontology (GO terms) from our gene-set analysis.

Finally, we performed a partitioned heritability analysis for the SNHL EUA MVP and UKB meta-analysis (N = 598,899 European subjects from MVP and UKB), partitioning SNPs into 3 groups: SNPs in protein coding genes (in HHLdb), SNPs in protein coding genes (not in HHLdb), and SNPs outside of protein coding genes (Table 2). SNP-based heritability estimates including 196 known HHL genes showed a 3.26-fold enrichment relative to all other protein-coding genes. However, the large majority of risk (99% of SNPs, explaining 97% of h^2^_SNP_) for polygenic SNHL is captured by SNPs outside of congenital HHL genes.

**Table 2.** Partitioned heritability estimates for sensorineural hearing loss GWAS (N = 598,899.

| **Category** | **SNPs** | **$h^2_{\text{SNP}}$** |
| --- | --- | --- |
| All SNPs | 1.00 | 1.00 |
| SNPs in protein coding genes (in HHLdb) | 0.01 | 0.03 |
| SNPs in protein coding genes (not in HHLdb) | 0.38 | 0.54 |
| SNPs outside of protein coding genes | 0.61 | 0.43 |

<sup>a</sup> Calculated as the proportion of $h^2_{\text{SNP}}$ accounted for by the category divided by the proportion of SNPs in the category. Abbreviations: HHLdb, Hereditary Hearing Loss Database; $h^2_{\text{SNP}}$ , SNP-based heritability

18,899 European subjects from MVP and UKB)
| SE | Enrichment<sup>a</sup> |
| --- | --- |
| 0.00 | 1.00 |
| 0.01 | 4.58 |
| 0.02 | 1.40 |
| 0.02 | 0.71 |

in of the genome-wide set of SNPs that were annotated to the category

## Discussion

### Million Veteran Program multi-ancestry analysis

This study introduces MVP hearing data based solely on a clinical diagnosis of sensorineural hearing loss. MVP GWAS alone elicits a total of 57 distinct loci with 76 genes in gene-based analysis and significant relevant otologic pathways implicated by gene-set analysis. We demonstrate that this phenotype approach is broadly successful for variant identification. A benefit of our method is the exclusion of many other diseases that lead to hearing loss, e.g. acoustic neuroma, otosclerosis, sudden hearing loss, etc., which may have distinct genetic architectures that can lead to inflation in the results.

In addition to a phenotypic difference between MVP and UKB, i.e., ICD versus self-report, the two populations have contrasting demographics, environmental exposures, and slightly different mean ages. Unlike the civilian population of the UKB, MVP is a group of ambulatory US Veterans consisting of greater than 90% males who have been exposed to noise levels during their military career^53^ of up to 126 dB^17^, as well as a 20% prevalence of head injury, another cause for hearing loss^54^. Although we cannot distinguish the effects of noise and head trauma from age in this study, the MVP cohort might have elicited the phenotypic expression of variants related to these differences.

Nevertheless, despite these dissimilarities, there appears to be a commonality of hearing loss genetic architecture between the MVP and UKB datasets where the studies estimated 3,624 (67.0%) shared causal variants. Importantly, this similarity indicates that the data are suitable for aggregation, and meta-analysis of the two identified 108 loci^12,16^, 54 with a novel prioritized and/or protein-coding gene, and a total of 100 significant novel genes in gene-analysis.

### Novel prioritized genes

Among our newly reported genes, at least five of the “top ten hits” within novel prioritized genes are worth noting as cochlear-related (Supplementary Table 10). *DPT* has been identified in familial Meniere’s disease, a syndrome consisting of episodic hearing loss, vertigo and tinnitus, and is an extracellular matrix protein that functions in cell matrix interactions and assembly^55^. *SETBP1* mutations are a cause of Schinzel-Giedion syndrome, characterized by distinct facial features, malformations, and SNHL^56^. *WNT7A* regulates axon outgrowth in the cochlea^57^ during inner ear development and is responsible for planarity of hair cell bundles^58^. *EVL* localizes at the cuticular plate supporting inner ear hair cells^59^. Expression of the transcription factor *BACH2*, under control of *miR-96* in the murine model, is altered in noise trauma, and plays a mediation role in oxidative stress^60^.

### Monogenic hearing loss genes and a plausible explanation of significant GWAS findings

Partitioned SNP-based heritability based on our results of known monogenic hearing loss genes showed a 3.26-fold enrichment relative to other protein-coding genes. This is in agreement with other studies tying hereditary hearing loss (HHL) genes to the polygenic disorder of adult SNHL, where polygenic risk scores from a self-reported hearing loss GWAS predicted higher odds of children’s hearing loss^61^. Separately, multiple studies of self-reported hearing difficulties have also identified an increased burden of rare variants within HHL genes^16,20,62–65^. Thus, while childhood hearing loss is known to be at least 50% monogenic in origin^66^, it appears that adult-onset SNHL has genetic signals in common with HHL.

To organize our results, it is helpful to examine the function of hereditary hearing loss genes and additional genes identified as significant in this study that are directly related to inner ear physiology (See Fig. 4), particularly within the context of the stereocilia of the inner and outer hair cells. Parallel actin fibers in stereocilia are critical for stiffness in response to the fluid wave. Within these structures, *ESPN* encodes an actin-binding protein required for assembly and stabilization^67^, *TMPRSS3* and *CLRN2* express stereociliary proteins critical for hair cell survival^68^, and *WNT7A* directs the uniform 3-dimensional polarity of outer hair cells.

In response to the fluid/sound wave, stereociliae of hair cells bend at their rootlets (Fig. 4b, 4c). *TRIOBP, CLIC5,* and *ATP2B2*, all non-syndromal congenital hearing loss genes, are expressed in this subcellular area^69–72^. *TRIOBP* wraps around actin providing stiffness and support, and *CLIC5* is a chloride channel protein that stabilizes the attachment of the stereocilia plasma membrane to its base^73–75^. *ATP2B2* expresses a transmembrane ATPase at the lower half of the stereocilia and cuticular plate, where it regulates Ca^++^ concentration within the cytosol. Although not on the HHL list, *SPTBN1* is expressed as part of the support structure at the area of the rootlet as well. *SPTBN1* is associated with an autosomal dominant neurodevelopmental disorder which includes SNHL^76^. In support of the “sensory mechanical stimulus pathway” (*p-* value 6.42E-09), our human-based transcription analysis of the organ of Corti indicates hair cell enrichment (Supplementary Table 15).

Bending at the rootlet allows for increased tension on tip-links, which in turn controls the opening of the mechanotransduction ion channel (MET). *CDH23*, prioritized by FLAME^38^, constitutes the upper half of the tip-link, and *MYO7A* tethers this link to an actin bundle within the stereocilium. These genes are responsible for different forms of Usher’s syndrome, the most common genetic cause of congenital deafness. In this critical area of the sensory apparatus, both *CLRN2* and *LOXHD1* are engaged in MET maintenance at the tip of the stereocilia^77^.

OHCs are tethered at their apex and base. At their apex, the tectorial membrane (TM) (Fig. 4b)^77^ is a gelatinous structure providing a static force to the shearing motion of the sound wave. *COL11A1* and *COL9A3* code for two α-chain collagens providing upper support within the tectorial membrane. They individually cause different variants of Stickler’s Syndrome^78^. Other significant support genes include *GAS2*, identified in pillar and Deiters’ cells as a microtubule regulatory gene, maintaining arrangement of the actin skeleton^79^.

In a separate micro-anatomical area, our human cochlear cell transcription analysis noted enrichment within melanocytes, located in the stria vascularis, and functioning to maintain the endocochlear potential. Although not on the HHL list, *ATP6V0A4,* identified within "sensory perception of mechanical stimulus", is a known hearing loss gene^80^ expressed in the basal stria cells, where it acts as a proton pump, maintaining the pH of endolymph bathing the apical region of the hair cell. *SOX10* (Waardenberg’s Syndrome), also found within the stria vascularis, expresses an aquaporin repressor^81^.

Associated with the nerve signal through the spiral ganglion, other significant congenital hearing loss genes were identified related to neurons, the ribbon synapse, and the myelin sheath (Fig. 4b). Studies of both age-related and noise-induced SNHL have found that after injury, up to 50% of ribbon synapses can be lost prior to an objective audiogram threshold change^82,83^ and this finding has been correlated to loss of speech intelligibility^84,85^. These synapses connect the IHC cells to Type I spiral ganglion nerve cells^86^. Located within this synaptic area are *LMX1A*, a neuro-restrictive silencer^87^ expressed in Type I spiral ganglion nerves. *QKI* is identified in a murine model as essential for proper myelination of spiral ganglion neurons and auditory nerve fibers^85^. In aggregate, these genes identified as significant in prioritization and gene-based analysis provide putative points of injury to the hearing sensory mechanism to be explored in future studies.

### Limitations

This study has limitations. Although an ICD diagnosis is a step towards an objective phenotype, it does not describe the state of individual audiogram frequencies, and heritability studies have indicated that a good portion of the heritability of SNHL is frequency-specific^88^. Further, while the genes identified provide biologically plausible explanations for SNHL, until now, expression data in the human cochlea has not been available. While this study used expression data from one human inner ear sample, further study for causative variants requires whole genome sequencing and multiple human cochlear methylation/expression studies.

### Conclusions

In GWAS of SNHL combining MVP and UKB, we have identified 108 loci containing 54 novel protein-coding genes and/or prioritized genes, 100 novel genes in gene-based analysis, and significant pathways related to the structure of the cochlea. Despite different phenotypes, demographics, and exposures we find substantial overlapping genetic signals. Although over 95% of the risk for this polygenic disorder is captured by SNPs outside of congenital HHL genes, SNP-based heritability estimates 3.26-fold enrichment of these genes relative to all other protein-coding genes. These findings will provide new targets of investigation of druggable genes and pathways for treatment of this pervasive and currently permanent condition. Our next tasks are to analyze genetic signals from individual audiogram frequencies within larger studies and search for phenotypes with increased heritability of this pervasive disorder.

## Data Availability

Summary statistics for MVP analyses will be deposited upon publication on dbGaP under accession number phs001672 (https://www.ncbi.nlm.nih.gov/projects/gap/cgi-bin/study.cgi?study_id=phs001672.v11.p1). MVP summary data access can be obtained by
submitting a data access request through dbGaP; raw data are protected and are not available due to privacy reasons. The codes for the analysis are available on Github (https://github.com/nievergeltlab/)

## Abbreviations

AFA: African Americans
AIC: Akaike information criterion
ANNOVAR: annotation variation software
BM: basement membrane
CADD: combined annotation dependent depletion
EUA: European ancestry
FUMA: functional mapping and annotation
GO: gene ontology
GWAS: genome-wide association study
GWS: genome-wide significant
*h_2_*SNP: heritability of single nucleotide polymorphisms
IAA: indigenous American ancestry
ICD: international classification of diseases
IHC: inner hair cells
LD: linkage disequilibrium
LDSC: linkage disequilibrium score regression
MAF: minor allele frequency
MAGMA: multi-marker analysis of genomic annotation
MET: mechanoelectrical transduction channels
MsigDB: molecular signatures database
MVP: Million Veteran Program
OC: Organ of Corti
OHC: outer hair cells
PC: principal component
SNHL: sensorineural hearing loss
SNP: single nucleotide polymorphism
TM: tectorial membrane
UKB: United Kingdom Biobank
YLD: years lived with disability

## Supplementary Information

**Additional file 1: Supplementary Table 1.** MVP demographics. **Supplementary Table 2.** Genome-wide significant loci in MVP across ancestries. **Supplementary Table 3.** Annotations for SNPs in LD with significant SNPs in the MVP multi-ancestry meta-analysis. **Supplementary Table 4.** Genome-wide significant genes from gene-based analyses across MVP ancestries. **Supplementary Table 5.** MAGMA gene-set analysis, including 17,012 pre-defined curated gene sets and GO terms obtained from MsigDB in the MVP multi-ancestry meta-analysis. **Supplementary Table 6.** Genes within the 3 significant gene-sets from the MAGMA gene-set analysis (MVP multi-ancestry meta-analysis). **Supplementary Table 7.** SNP-based heritability and genetic correlation across of MVP and UKB EUA data. **Supplementary Table 8.** MiXeR analysis of SNHL GWAS in European ancestry data. **Supplementary Table 9.** Genome-wide significant loci in the European ancestry meta-analysis (MVP EUA and UKB). **Supplementary Table 10.** Extended Table 1: Genome-wide significant loci in meta-analysis of sensorineural hearing loss (N=725,904 MVP multi-ancestry and UKB individuals). **Supplementary Table 11.** Annotations for all SNPs in LD with significant SNPs in the MVP multi-ancestry and UKB meta-analysis. **Supplementary Table 12.** Gene-based analyses in the MVP multi-ancestry and UKB cohorts. **Supplementary Table 13.** MAGMA gene-set analysis, including 17009 pre-defined curated gene sets and GO terms obtained from MsigDB in the MVP multi-ancestry and UKB EUA meta-analysis. **Supplementary Table 14.** Genes within the 3 significant gene-sets from the MAGMA gene-set analysis (MVP multi-ancestry and UKB meta-analysis). **Supplementary Table 15.** Tissue enrichment analysis for 31 and 71 tissue types across the European ancestry MVP and UKB cohorts. **Supplementary Table 16**. Cell type enrichment analyses in cochlear cells based on mouse gene expression data in MVP EUA and UKB. Supplementary Table 17. Hereditary Hearing Loss genes implicated across analyses (HereditaryHearingLoss.org database)

**Additional file 2: Supplementary Figure 1.** Ancestry-stratified GWAS and meta-analysis for SNHL in the MVP. **Supplementary Figure 2.** Ancestry-stratified gene-based analyses of SNHL in the MVP. **Supplementary Figure 3.** Gene-based analyses of the MVP multi-ancestry meta-analysis and the UKB SNHL GWAS. **Supplementary Figure 4.** Gene-tissue expression analyses of SNHL. Supplementary Figure 5. Cochlear cell type enrichment analyses based on mouse data.

**Additional file 3:** MVP Core Acknowledgements.

## Acknowledgements

We thank all the participants in the UK Biobank and the Million Veteran Program. This research is based on data from the Million Veterans Program, Office of Research and Development, Veterans Health Administration, and was supported by MVP000 as well as awards #I01RX004293 and #I01BX005920. This publication does not represent the views of the Department of Veteran Affairs or the United States Government. See MVP Core Acknowledgements in Additional file 3.

## Authors’ contributions

RC, AXM, and CMN designed the study. AXM, EAM, and CMN conducted GWAS data analysis. RC, CEM, JJ, EAM, AXM, and CMN performed post-GWAS analysis. RC, JJ, CEM, and EAM visualized the data. All of the authors participated in the interpretation of data and revision of the manuscript for intellectual content. RC and CMN obtained primary funding. RC supervised the phenotype and CMN supervised the statistical analysis of GWAS results. All authors read and approved of the final manuscript.

## Availability of data and materials

Summary statistics for MVP analyses will be deposited upon publication on dbGaP under accession number phs001672 (https://www.ncbi.nlm.nih.gov/projects/gap/cgi-bin/study.cgi?study_id=phs001672.v11.p1). MVP summary data access can be obtained by submitting a data access request through dbGaP; raw data are protected and are not available due to privacy reasons.

The codes for the analysis are available on Github (https://github.com/nievergeltlab/)

## Declarations

### Ethics approval and consent to participate

The use of MVP individual-level data was conducted under project #MVP039 and #MVP060. MVP is approved by the VA Central IRB, and the projects were also approved by local VA IRB in San Diego. This research was performed using the UKB Resource under application 40951. UKB has approval from the North West Multi-center Research Ethics Committee (MREC) as a Research Tissue Bank (RTB) approval. This approval means that separate ethical approval is waived and that researchers can operate under the RTB approval.

### Consent for publication

This manuscript was approved for submission and publication by the MVP Publications Committee.

## References

1. Nocini, R., Henry, B.M., Lippi, G. & Mattiuzzi, C. Estimating the worldwide burden of health loss due to hearing loss. Eur J Public Health 33, 146–148 (2023). PMID 36377968

2. Dawes, P. et al. Hearing loss and cognition: the role of hearing AIDS, social isolation and depression. PLoS One 10, e0119616 (2015). PMID 25760329

3. Yévenes-Briones, H. et al. Association Between Hearing Loss and Impaired Physical Function, Frailty, and Disability in Older Adults: A Cross-sectional Study. JAMA Otolaryngol Head Neck Surg 147, 951–958 (2021). PMID 34554203

4. Jafari, Z., Kolb, B.E. & Mohajerani, M.H. Age-related hearing loss and tinnitus, dementia risk, and auditory amplification outcomes. Ageing Res Rev 56, 100963 (2019). PMID 31557539

5. Dobie, R.A. The burdens of age-related and occupational noise-induced hearing loss in the United States. Ear Hear 29, 565–77 (2008). PMID 18469718

6. Duan, H. et al. Heritability of Age-Related Hearing Loss in Middle-Aged and Elderly Chinese: A Population-Based Twin Study. Ear Hear 40, 253–259 (2019). PMID 29794565

7. Bogo, R. et al. The role of genetic factors for hearing deterioration across 20 years: a twin study. J Gerontol A Biol Sci Med Sci 70, 647–53 (2015). PMID 25665831

8. Walls WD, A.H., Smith RJH. Hereditary Hearing Loss Homepage.

9. Fransen, E. et al. Genome-wide association analysis demonstrates the highly polygenic character of age-related hearing impairment. Eur J Hum Genet 23, 110–5 (2015). PMID 24939585

10. De Angelis, F. et al. Sex differences in the polygenic architecture of hearing problems in adults. Genome Med 15, 36 (2023). PMID 37165447

11. Jiang, L., Wang, D., He, Y. & Shu, Y. Advances in gene therapy hold promise for treating hereditary hearing loss. Mol Ther 31, 934–950 (2023). PMID 36755494

12. Wells, H.R.R. et al. GWAS Identifies 44 Independent Associated Genomic Loci for Self-Reported Adult Hearing Difficulty in UK Biobank. Am J Hum Genet 105, 788–802 (2019). PMID 31564434

13. McCullagh, M.C., Raymond, D., Kerr, M.J. & Lusk, S.L. Prevalence of hearing loss and accuracy of self-report among factory workers. Noise Health 13, 340–7 (2011). PMID 21959114

14. Brennan-Jones, C.G. et al. Self-reported hearing loss and manual audiometry: A rural versus urban comparison. Aust J Rural Health 24, 130–5 (2016). PMID 26311193

15. Cherny, S.S. et al. Self-reported hearing loss questions provide a good measure for genetic studies: a polygenic risk score analysis from UK Biobank. Eur J Hum Genet 28, 1056–1065 (2020). PMID 32203203

16. Trpchevska, N. et al. Genome-wide association meta-analysis identifies 48 risk variants and highlights the role of the stria vascularis in hearing loss. Am J Hum Genet 109, 1077–1091 (2022). PMID 35580588

17. Jokel, C., Yankaskas, K. & Robinette, M.B. Noise of military weapons, ground vehicles, planes and ships. J Acoust Soc Am 146, 3832 (2019). PMID 31795677

18. Praveen, K. et al. Population-scale analysis of common and rare genetic variation associated with hearing loss in adults. Commun Biol 5, 540 (2022). PMID 35661827

19. Naderi, E. et al. The genetic contribution of the X chromosome in age-related hearing loss. Front Genet 14, 1106328 (2023). PMID 36896235

20. Ivarsdottir, E.V. et al. The genetic architecture of age-related hearing impairment revealed by genome-wide association analysis. Commun Biol 4, 706 (2021). PMID 34108613

21. Liu, W., Johansson, A., Rask-Andersen, H. & Rask-Andersen, M. A combined genome-wide association and molecular study of age-related hearing loss in H. sapiens. BMC Med 19, 302 (2021). PMID 34847940

22. Kalra, G. et al. Biological insights from multi-omic analysis of 31 genomic risk loci for adult hearing difficulty. PLoS Genet 16, e1009025 (2020). PMID 32986727

23. Gaziano, J.M. et al. Million Veteran Program: A mega-biobank to study genetic influences on health and disease. J Clin Epidemiol 70, 214–23 (2016). PMID 26441289

24. Loh, P.R., Palamara, P.F. & Price, A.L. Fast and accurate long-range phasing in a UK Biobank cohort. Nat Genet 48, 811–6 (2016). PMID 27270109

25. Das, S. et al. Next-generation genotype imputation service and methods. Nat Genet 48, 1284–1287 (2016). PMID 27571263

26. McCarthy, S. et al. A reference panel of 64,976 haplotypes for genotype imputation. Nat Genet 48, 1279–83 (2016). PMID 27548312

27. Chen, C.Y. et al. Improved ancestry inference using weights from external reference panels. Bioinformatics 29, 1399–406 (2013). PMID 23539302

28. Nievergelt, C.M. et al. International meta-analysis of PTSD genome-wide association studies identifies sex- and ancestry-specific genetic risk loci. Nat Commun 10, 4558 (2019). PMID 31594949

29. Chang, C.C. et al. Second-generation PLINK: rising to the challenge of larger and richer datasets. Gigascience 4, 7 (2015). PMID 25722852

30. Manichaikul, A. et al. Robust relationship inference in genome-wide association studies. Bioinformatics 26, 2867–73 (2010). PMID 20926424

31. Abraham, G., Qiu, Y. & Inouye, M. FlashPCA2: principal component analysis of Biobank-scale genotype datasets. Bioinformatics 33, 2776–2778 (2017). PMID 28475694

32. Cook, J.P., Mahajan, A. & Morris, A.P. Guidance for the utility of linear models in meta-analysis of genetic association studies of binary phenotypes. Eur J Hum Genet 25, 240–245 (2017). PMID 27848946

33. Willer, C.J., Li, Y. & Abecasis, G.R. METAL: fast and efficient meta-analysis of genomewide association scans. Bioinformatics 26, 2190–1 (2010). PMID 20616382

34. Watanabe, K., Taskesen, E., van Bochoven, A. & Posthuma, D. Functional mapping and annotation of genetic associations with FUMA. Nat Commun 8, 1826 (2017). PMID 29184056

35. Wang, K., Li, M. & Hakonarson, H. ANNOVAR: functional annotation of genetic variants from high-throughput sequencing data. Nucleic Acids Res 38, e164 (2010). PMID 20601685

36. Rentzsch, P., Witten, D., Cooper, G.M., Shendure, J. & Kircher, M. CADD: predicting the deleteriousness of variants throughout the human genome. Nucleic Acids Res 47, D886–D894 (2019). PMID 30371827

37. Dong, S. et al. Annotating and prioritizing human non-coding variants with RegulomeDB v.2. Nat Genet 55, 724–726 (2023). PMID 37173523

38. Schipper, M. et al. Prioritizing effector genes at trait-associated loci using multimodal evidence. Nat Genet 57, 323–333 (2025). PMID 39930082

39. Hutchinson, A., Watson, H. & Wallace, C. Improving the coverage of credible sets in Bayesian genetic fine-mapping. PLoS Comput Biol 16, e1007829 (2020). PMID 32282791

40. Weeks, E.M. et al. Leveraging polygenic enrichments of gene features to predict genes underlying complex traits and diseases. Nat Genet 55, 1267–1276 (2023). PMID 37443254

41. Pruim, R.J. et al. LocusZoom: regional visualization of genome-wide association scan results. Bioinformatics 26, 2336–7 (2010). PMID 20634204

42. de Leeuw, C.A., Mooij, J.M., Heskes, T. & Posthuma, D. MAGMA: generalized gene-set analysis of GWAS data. PLoS Comput Biol 11, e1004219 (2015). PMID 25885710

43. Liberzon, A. et al. The Molecular Signatures Database (MSigDB) hallmark gene set collection. Cell Syst 1, 417–425 (2015). PMID 26771021

44. van der Valk, W.H. et al. A single-cell level comparison of human inner ear organoids with the human cochlea and vestibular organs. Cell Rep 42, 112623 (2023). PMID 37289589

45. Jean, P. et al. Single-cell transcriptomic profiling of the mouse cochlea: An atlas for targeted therapies. Proc Natl Acad Sci U S A 120, e2221744120 (2023). PMID 37339214

46. Hoa, M. et al. Characterizing Adult Cochlear Supporting Cell Transcriptional Diversity Using Single-Cell RNA-Seq: Validation in the Adult Mouse and Translational Implications for the Adult Human Cochlea. Front Mol Neurosci 13, 13 (2020). PMID 32116546

47. Blake, J.A. et al. Mouse Genome Database (MGD): Knowledgebase for mouse-human comparative biology. Nucleic Acids Res 49, D981–D987 (2021). PMID 33231642

48. Bulik-Sullivan, B.K. et al. LD Score regression distinguishes confounding from polygenicity in genome-wide association studies. Nat Genet 47, 291–5 (2015). PMID 25642630

49. Lee, S.H., Wray, N.R., Goddard, M.E. & Visscher, P.M. Estimating missing heritability for disease from genome-wide association studies. Am J Hum Genet 88, 294–305 (2011). PMID 21376301

50. Holland, D. et al. Beyond SNP heritability: Polygenicity and discoverability of phenotypes estimated with a univariate Gaussian mixture model. PLoS Genet 16, e1008612 (2020). PMID 32427991

51. Frei, O. et al. Bivariate causal mixture model quantifies polygenic overlap between complex traits beyond genetic correlation. Nat Commun 10, 2417 (2019). PMID 31160569

52. Johnson, K.R., Zheng, Q.Y. & Noben-Trauth, K. Strain background effects and genetic modifiers of hearing in mice. Brain Res 1091, 79–88 (2006). PMID 16579977

53. Yankaskas, K. Prelude: noise-induced tinnitus and hearing loss in the military. Hear Res 295, 3–8 (2013). PMID 22575206

54. Chen, J.X. et al. Systematic review of hearing loss after traumatic brain injury without associated temporal bone fracture. Am J Otolaryngol 39, 338–344 (2018). PMID 29506762

55. Escalera-Balsera, A., Roman-Naranjo, P. & Lopez-Escamez, J.A. Systematic Review of Sequencing Studies and Gene Expression Profiling in Familial Meniere Disease. Genes (Basel) 11(2020). PMID 33260921

56. Hoischen, A. et al. De novo mutations of SETBP1 cause Schinzel-Giedion syndrome. Nat Genet 42, 483–5 (2010). PMID 20436468

57. Munnamalai, V. & Fekete, D.M. Wnt signaling during cochlear development. Semin Cell Dev Biol 24, 480–9 (2013). PMID 23548730

58. Dabdoub, A. et al. Wnt signaling mediates reorientation of outer hair cell stereociliary bundles in the mammalian cochlea. Development 130, 2375–84 (2003). PMID 12702652

59. Yan, K. et al. EVL is not essential for cuticular plate and stereocilia development in mouse auditory hair cells. FEBS Lett 599, 330–339 (2025). PMID 39300480

60. Xia, R. et al. Therapeutic restoration of miR-96 prevents hearing loss in mice through modulation of noise-induced and genetic pathways. iScience 28, 112355 (2025). PMID 40641557

61. Wang, J. et al. Polygenic Risk Scores and Hearing Loss Phenotypes in Children. JAMA Otolaryngol Head Neck Surg (2024). PMID 39509092

62. Hui, D. et al. Gene burden analysis identifies genes associated with increased risk and severity of adult-onset hearing loss in a diverse hospital-based cohort. PLoS Genet 19, e1010584 (2023). PMID 36656851

63. Lewis, M.A. et al. Whole exome sequencing in adult-onset hearing loss reveals a high load of predicted pathogenic variants in known deafness-associated genes and identifies new candidate genes. BMC Med Genomics 11, 77 (2018). PMID 30180840

64. Lewis, M.A., Schulte, B.A., Dubno, J.R. & Steel, K.P. Investigating the characteristics of genes and variants associated with self-reported hearing difficulty in older adults in the UK Biobank. BMC Biol 20, 150 (2022). PMID 35761239

65. Cornejo-Sanchez, D.M. et al. Rare-variant association analysis reveals known and new age-related hearing loss genes. Eur J Hum Genet 31, 638–647 (2023). PMID 36788145

66. Khela, H. & Kenna, M.A. Genetics of pediatric hearing loss: A functional perspective. Laryngoscope Investig Otolaryngol 5, 511–519 (2020). PMID 32596495

67. Sekerkova, G., Zheng, L., Loomis, P.A., Mugnaini, E. & Bartles, J.R. Espins and the actin cytoskeleton of hair cell stereocilia and sensory cell microvilli. Cell Mol Life Sci 63, 2329–41 (2006). PMID 16909209

68. Dunbar, L.A. et al. Clarin-2 is essential for hearing by maintaining stereocilia integrity and function. EMBO Mol Med 11, e10288 (2019). PMID 31448880

69. Katsuno, T. et al. TRIOBP-5 sculpts stereocilia rootlets and stiffens supporting cells enabling hearing. JCI Insight 4(2019). PMID 31217345

70. Fasquelle, L. et al. Tmprss3, a transmembrane serine protease deficient in human DFNB8/10 deafness, is critical for cochlear hair cell survival at the onset of hearing. J Biol Chem 286, 17383–97 (2011). PMID 21454591

71. Liu, W. et al. Expression of trans-membrane serine protease 3 (TMPRSS3) in the human organ of Corti. Cell Tissue Res 372, 445–456 (2018). PMID 29460002

72. Pacentine, I., Chatterjee, P. & Barr-Gillespie, P.G. Stereocilia Rootlets: Actin-Based Structures That Are Essential for Structural Stability of the Hair Bundle. Int J Mol Sci 21(2020). PMID 31947734

73. Cooper, N.P., Vavakou, A. & van der Heijden, M. Vibration hotspots reveal longitudinal funneling of sound-evoked motion in the mammalian cochlea. Nat Commun 9, 3054 (2018). PMID 30076297

74. Gagnon, L.H. et al. The chloride intracellular channel protein CLIC5 is expressed at high levels in hair cell stereocilia and is essential for normal inner ear function. J Neurosci 26, 10188–98 (2006). PMID 17021174

75. Silverstein, R.S. & Tempel, B.L. Atp2b2, encoding plasma membrane Ca2+-ATPase type 2, (PMCA2) exhibits tissue-specific first exon usage in hair cells, neurons, and mammary glands of mice. Neuroscience 141, 245–57 (2006). PMID 16675132

76. Cousin, M.A. et al. Pathogenic SPTBN1 variants cause an autosomal dominant neurodevelopmental syndrome. Nat Genet 53, 1006–1021 (2021). PMID 34211179

77. Wang, P. et al. LOXHD1 is indispensable for coupling auditory mechanosensitive channels to the site of force transmission. Res Sq (2024). PMID 38260480

78. Shpargel, K.B., Makishima, T. & Griffith, A.J. Col11a1 and Col11a2 mRNA expression in the developing mouse cochlea: implications for the correlation of hearing loss phenotype with mutant type XI collagen genotype. Acta Otolaryngol 124, 242–8 (2004). PMID 15141750

79. Chen, T. et al. Cochlear supporting cells require GAS2 for cytoskeletal architecture and hearing. Dev Cell 56, 1526–1540 e7 (2021). PMID 33964205

80. Stover, E.H. et al. Novel ATP6V1B1 and ATP6V0A4 mutations in autosomal recessive distal renal tubular acidosis with new evidence for hearing loss. J Med Genet 39, 796–803 (2002). PMID 12414817

81. Szeto, I.Y.Y. et al. SOX9 and SOX10 control fluid homeostasis in the inner ear for hearing through independent and cooperative mechanisms. Proc Natl Acad Sci U S A 119, e2122121119 (2022). PMID 36343245

82. Fernandez, K.A. et al. Noise-induced Cochlear Synaptopathy with and Without Sensory Cell Loss. Neuroscience 427, 43–57 (2020). PMID 31887361

83. Kujawa, S.G. & Liberman, M.C. Synaptopathy in the noise-exposed and aging cochlea: Primary neural degeneration in acquired sensorineural hearing loss. Hear Res 330, 191–9 (2015). PMID 25769437

84. Bramhall, N.F. & McMillan, G.P. Perceptual Consequences of Cochlear Deafferentation in Humans. Trends Hear 28, 23312165241239541 (2024). PMID 38738337

85. Panganiban, C.H. et al. Noise-Induced Dysregulation of Quaking RNA Binding Proteins Contributes to Auditory Nerve Demyelination and Hearing Loss. J Neurosci 38, 2551–2568 (2018). PMID 29437856

86. Jean, P., Ozcete, O.D., Tarchini, B. & Moser, T. Intrinsic planar polarity mechanisms influence the position-dependent regulation of synapse properties in inner hair cells. Proc Natl Acad Sci U S A 116, 9084–9093 (2019). PMID 30975754

87. Aoki, H., Hara, A. & Kunisada, T. White spotting phenotype induced by targeted REST disruption during neural crest specification to a melanocyte cell lineage. Genes Cells 20, 439–49 (2015). PMID 25818501

88. Wingfield, A. et al. A twin-study of genetic contributions to hearing acuity in late middle age. J Gerontol A Biol Sci Med Sci 62, 1294–9 (2007). PMID 18000151

