## Additional file 2 for "Genome-wide association study meta-analysis brings monogenic hearing loss genes into the polygenic realm"

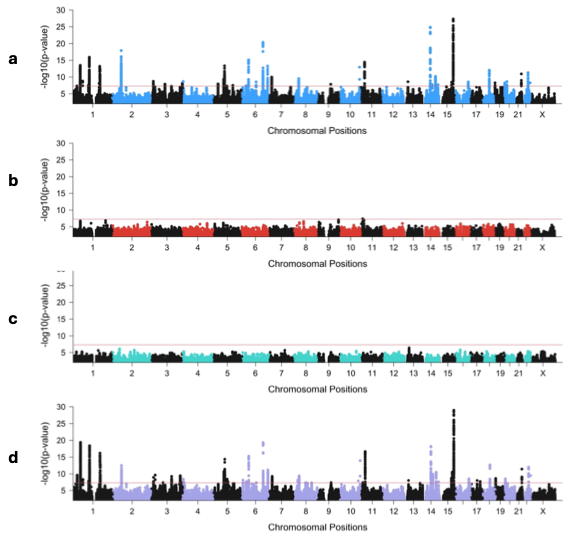

**Supplementary Figure 1. Ancestry-stratified GWAS and meta-analysis for SNHL in the MVP.**

Manhattan plots of sensorineural hearing loss (SNHL) GWAS conducted within each ancestry strata of the MVP and the multi-ancestry meta-analysis. The red line represents genome-wide significance (GWS) at p < 5 × 10−8. **a)** European ancestry (EUA) GWAS of 176,393 cases and 172,117 controls identified 43 GWS loci. **b)** African ancestry (AFA) GWAS of 22,757 cases and 74,842 controls identified 1 significant locus. **c)** Indigenous American (IAA) ancestry GWAS of 11,090 cases, 18,316 controls. **d)** Meta-analysis of the three ancestries identified 52 GWS loci.

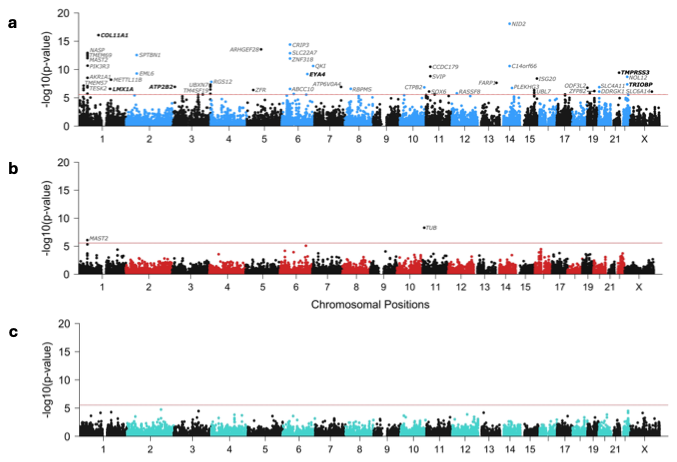

**Supplementary Figure 2. Ancestry-stratified gene-based analyses of SNHL in the MVP.**

Manhattan plots of gene-based analysis of sensorineural hearing loss (SNHL) conducted within each ancestry strata of the MVP. The red line indicates the gene-wide significance threshold at p < 2.66 × 10−6 (Bonferroni correction for 18,809 genes tested). A selection of significant genes have been labeled. Significant genes that are also listed in HereditaryHearingLoss.org have been labeled in bold font. a) European ancestry (EUA) gene-based analysis of 176,393 cases and 172,117 controls identified 62 significant genes. b) African ancestry (AFA) gene-based analysis of 22,757 cases and 74,842 controls identified 2 significant genes. c) Indigenous American ancestry (IAA) gene-based analysis of 11,090 cases, 18,316 controls.

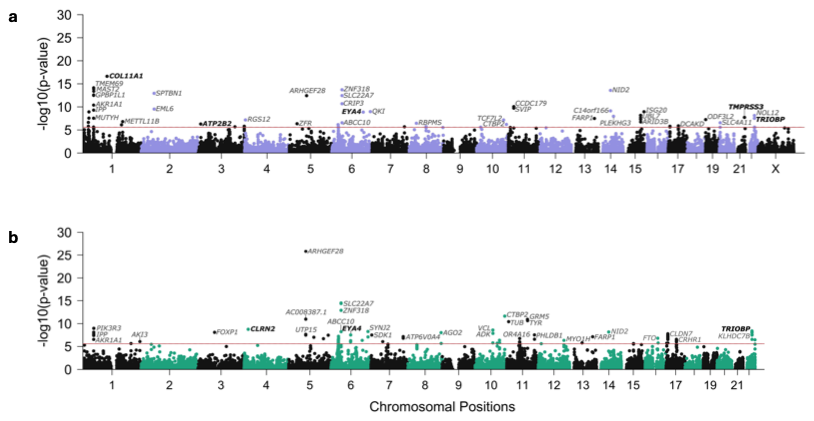

**Supplementary Figure 3. Gene-based analyses of the MVP multi-ancestry meta-analysis and the UKB SNHL GWAS.**

Manhattan plots of gene-based analysis of sensorineural hearing loss (SNHL). The red line indicates the gene-wide significance threshold at p < 2.66 × 10−6 (Bonferroni correction for 18,809 genes tested). A selection of significant genes have been labeled. Significant genes that are also listed in HereditaryHearingLoss.org have been labeled in bold. **a)** Gene-based analysis of the MVP multi-ancestry meta-analysis (210,240 cases and 265,275 controls) identified 64 significant genes. **b)** UKB gene-based analysis of 87,056 cases and 163,333 controls identified 97 significant genes. The X chromosome was not included in the UKB analysis.

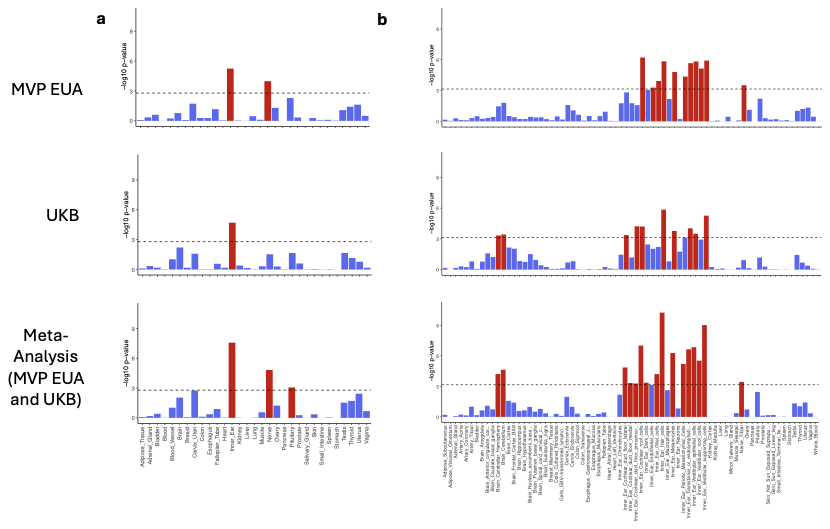

**Supplementary Figure 4. Supplementary Figure 4. Gene-tissue expression analyses of SNHL based on human data.**

MAGMA gene-tissue expression analyses for adult inner ear and GTEx v8 tissues in sensorineural hearing loss (SNHL), conducted in the MVP European ancestry (EUA), UKB, and meta-analysis of the MVP EUA and UKB. Bars denote -log10 p-values. Dotted lines indicate significance after Bonferroni-adjustment for the number of tissues tested (panel a, p < 1.61 × 10−3 for 31 general tissue types tested; panel b, p < 7.04 × 10−4 for 71 specific tissues tested). Bonferroni significant tissue associations are also indicated through red shading of bars. **a)** Gene-tissue analysis of 31 general tissue types indicates a significant enrichment of the inner ear in all three analyses, as well as the nerve in MVP EUA, and both the nerve and pituitary in the meta-analysis. **b)** Among the 71 specific tissues, 13 cochlear and vestibular cell types were significant in the meta-analysis, including 8 in UKB and 10 in MVP EUA. The cerebellum and cerebellar hemisphere are significant in both UKB and the meta-analysis (note: cerebellar hemisphere and cerebellum are the same tissue, with different RNA preservation after death).

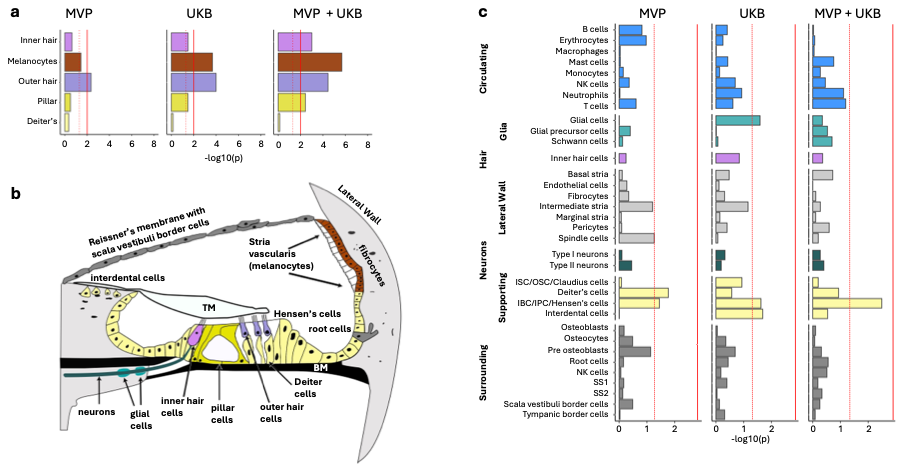

**Supplementary Figure 5. Cochlear cell type enrichment analyses analyses based on mouse data.**

**a)** Organ of Corti cell type enrichment analyses based on mouse data (Hoa et al. 2020). The bar chart depicts results of conditional MAGMA gene-property analyses for five different cell types. **b)** Cross-section of the Organ of Corti. The Organ of Corti is supported by the basilar membrane (BM) below and movement is restricted by gelatinous tectorial membrane (TM) above. c) Cochlear cell type enrichment analyses based on mouse data (Jean et al. 2023) depicts results of conditional MAGMA gene-property analyses for 34 different cell types. Bars denote -log10 p-values. Dotted line indicates p < 0.05. Solid line indicates significance after Bonferroni-adjustment for multiple comparisons (panel a, p < 0.01 for 5 cell types tested; panel c, p < 1.47 × 10−3 for 34 cell types tested). **Abbreviations:** ISC, inner sulcus cells; OSC, outer sulcus cells; IBC, inner border cells; IPC, inner phalangeal cells
